# Single-cell profiling reveals the sustained immune infiltration, surveillance, and tumor heterogeneity of infiltrative BCC

**DOI:** 10.1101/2022.08.22.22278093

**Authors:** Lingjuan Huang, Xianggui Wang, Shiyao Pei, Xin Li, Liang Dong, Xiaohui Bian, Hongyin Sun, Liping Jin, Huihui Hou, Wensheng Shi, Xiyuan Zhang, Lining Zhang, Shuang Zhao, Xiang Chen, Mingzhu Yin

## Abstract

**Background:** Infiltrative basal cell carcinoma (iBCC) is a particularly aggressive subtype of BCC that tends to recur after surgery, and its progression and malignancy are closely related to its interaction with the local tumor microenvironment (TME).

**Methods:** Here, we performed single-cell RNA sequencing (scRNA-seq) from 5 patients to determine the dynamic changes of TME between iBCC and adjacent normal skin (ANS).

**Results:** We found active immune collaborations among CXCL13^+^ follicular helper-like T cells (Tfh-like cells), SPP1^+^CXCL9/10^high^ Macro1, and plasma cells, which were enriched in iBCC. Specifically, SPP1^+^CXCL9/10^high^ Macro1 could activate plasma cells by BAFF signaling, and Tfh-like cells potentially recruited B/Plasma cells through CXCL13. The proinflammatory SPP1^+^CXCL9/10^high^ Macro1 and angiogenesis-related SPP1^+^CCL2^high^ Macro1 were characterized, revealing their heterogeneous phenotype within the TME. We also discovered a novel iBCC-enriched ANGPT2^+^ lymphatic endothelial cell (LEC) subtype with strong abilities to promote leukocyte migration and activation. Interestingly, we found upregulation of MHC-I molecules in fibroblasts in iBCC compared to those in ANS. However, we found that MDK signals derived from malignant basal cells (MBCs) were markedly increased, and their expression was an independent factor in predicting the infiltration depth of iBCC, emphasizing its role in driving malignancy and remodeling the TME. Additionally, we identified differentiation-associated SOSTDC1^+^IGFBP5^+^CTSV^+^ MB1 and epithelial mesenchyme transition (EMT)-associated TNC^+^SFRP1^+^CHGA^+^ MB2. The components of the two heterogeneous subpopulations in the TME might be effective predictors of the malignancy and prognosis of iBCC.

**Conclusions:** Altogether, our study is beneficial for understanding the cellular heterogeneity involved in the pathogenesis of iBCC and provides potential therapeutic targets for clinical research.

## Background

BCC is the most common cancer worldwide and accounts for 75% of nonmelanoma skin cancers[1]. BCC is derived from epidermal cells and often occurs on the face, damaging the aesthetics of patients and causing psychological burdens[2]. Ultraviolet (UV) exposure is the most important risk factor for BCC, and light-skinned people are also susceptible[3]. The main genetic mutations of BCC occur in the Hedgehog family (∼80%) and TP53 (∼60%)[4]. Genomic research has shown that BCC exhibits one of the highest tumor mutation burdens (TMB, 50–75 mutations/Mb) among malignancies[5]. Surgical resection is the curative option for most patients; however, some patients still develop postoperative recurrence that even leads to death[6]. Infiltrative BCC (iBCC), one of the clinical subtypes of BCC, has a tendency of destructive growth and invasion of surrounding tissue and has a higher risk of relapse after surgery[2]. Moreover, a small percentage could progress into unresectable advanced BCC, requiring systemic therapy[7]. Thus, there is an urgent need to comprehensively characterize the pathogenesis of iBCC to prevent its progression and alleviate the issue of recurrence.

The TME contains tumor and immune cells, a complicated network of stromal cells, and noncellular components, which are crucial for the initiation and progression of malignancies[8, 9]. In general, the microenvironment of tumors in the early stage tends to exhibit antitumor functions; however, once tumors develop into the late stage, they can transform into protumor functions[9]. Single-cell transcriptome sequencing (scRNA-seq) technology has led to tremendous breakthroughs in understanding the TME, such as intratumoral heterogeneity, specific transcriptome programs associated with tumor prognosis[10], and alterations in immune cellular collaboration[11, 12]. Yost et al. performed a dedicated scRNA-seq analysis of the change in T cells before and after immunotherapy in advanced BCC. However, a systemic single-cell atlas of iBCC in the early stage is lacking. The immune states, potential malignancy-driver molecules, relapse-associated mechanisms, and tumor cell heterogeneity of iBCC are largely unknown.

To fill this gap, we performed scRNA-seq of iBCC and adjacent normal skin (ANS) from 5 patients to comprehensively understand the characteristics and alterations of the TME in iBCC. Interestingly, we discovered active immune collaborations among tumor-enriched CD4^+^CXCL13^+^ T cells, SPP1^+^ CXCL9/10^high^ Macro1, and plasma cells and a novel-identified ANGPT2^high^ LEC subtype. The following two subtypes of proinflammatory SPP1+ Macro1 with distinct functions were identified. SPP1+CCL2^high^ Macro1, which was characterized by a higher angiogenic ability, and SPP1^+^ CXCL9/10^high^ Macro1, which was characterized by a T cell chemotaxis function, indicating the inner heterogeneous response and differentiation tendency of SPP1^+^ macrophages under TME stimulation. We also show that MBCs could remodel the TME by secreting the MDK signal, whose expression was associated with the infiltration depth of iBCC. Moreover, differentiation-associated SOSTDC1^+^IGFBP5^+^CTSV^+^ MB1 and EMT-associated TNC^+^SFRP1^+^CHGA^+^ MB2 were identified as two molecular types of iBCC, and their relative components within the TME may predict the malignancy level of iBCC. The MDK and two molecule types of iBCC identified in our study may serve as potential clinical predictors of the progression and relapse of iBCC.

## Methods

### Sample collection and patient clinical information

Tumor samples (n=5) and adjacent normal samples (n=3) were collected from 5 patients with diagnosed with invasive BCC. All five patients were from Xiangya Hospital of Central South University and signed an informed consent form for providing samples. Detailed clinical characteristics and pathological information are summarized in Table S1. Our study was approved by the institutional ethics committe of Xiangya Hospital of Central South University (2022020109).

### Single-cell isolation

Fresh clinical samples were collected after surgery and stored at 2–8 °C in MACS Tissue Storege Solution (Miltenyi Biotec). Firstly, sample tissues were minced into small pieces and then they were digested in GEXSCOPE Tissue Dissociation Solution (Singleron Biotechnologies) at 37 °C for 25 min with continuous agitation. After digestion, these suspended cells were washed by Dulbecco’s Phosphate Buffered Saline (DPBS, BI) and were sieved through a 40-micron sterile strainer (Falcon). The cells were then centrifuged at 2000 rpm for 10 min and the cell pellets were resuspended by appropriate buffer. Finally, the suspensions were stained with trypan blue (Sigma, United States) and the cellular viability was evaluated microscopically.

### Constructing single-cell libraries and sequencing

Two scRNA-seq platforms, including the GEXSCOPE and the BD Rhapsody platforms, were applied to construct single-cell libraries. The corresponding platforms of each sample were listed in Table S1. Detailed procedures are described in the following.

For the GEXSCOPE platform, the concentration of single-cell suspension was adjusted to 1×10^5^ cells/ml in DPBS (BI). A total of 170ul single-cell suspension were then loaded into microfluidic devices (GEXSCOPE Single Cell RNA-seq Kit, Singleron Biotechnologies). The subsequent procedures were performed according to the manufacturer’s protocol (Singleron Biotechnologies)[13]. Finally, the scRNA-seq libraries were sequenced on Illumina HiSeq X with 150 bp paired end reads.

For the BD Rhapsody platform, 400ul single-cell suspension containing around 4 × 10^4^ cells with cold Sample Buffer (BD Biosciences) were required. Then it was loaded onto a BD Rhapsody cartridge with >200,000 microwells. The following steps were performed according to the protocol of BD Rhapsody^TM^ Single cell 3’ whole transcriptome amplification (WTA) with sample multiplexing kit (BD Biosciences). Sequencing of the finished libraries was performed on Illumina HiSeq X with 150 bp paired end reads.

### Sequencing data preprocessing

For data generated with the Singleron, reads were mapped to the human genome (GRCh38) using scopetools (https://github.com/SingleronBio/SCOPE-tools). First, cell barcodes and unique molecular identifiers (UMIs) were extracted after filtering read one based on sequence information. Then the corrected barcodes and original UMI sequence were added to the ID of read two. Subsequently, read two were trimmed by cutadapt and then aligned to the reference genome with STAR [14, 15]. Furthermore, featureCounts was used to target reads to the genomic position of genes (ensemble version 99)[16]. Finally, reads with the same cell barcode, UMI and gene were grouped together and the number of UMI per gene were calculated for each cell.

For data generated with BD Rhapsody, the FASTQ files were processed according to the BD Rhapsody Analysis pipeline (BD Biosciences), which was achieved in the CWL-runner. In brief, low-quality read pairs were filtered and the quality-filtered R1 reads were analyzed to identify cell label and UMI sequences. After this, the filtered R2 reads were mapped to the transcriptome by STAR in the pipline. Reads with the same cell label, the same UMI sequence and the same gene were collapsed into a single raw molecule. The obtained counts were adjusted by BD Biosciences-developed error correction algorithm—recursive substitution error correction (RSEC) to correct sequencing and PCR errors. Barcoded oligo-conjugated antibodies (single-cell multiplexing kit; BD Biosciences) were adapted to speculate the origin of sample.

### Quality control, dimensionality reduction, clustering

The UMI count data of each sample was imported into Seurat to process and analyze (V4.1.0). Briefly, each sample dataset was converted to a Seurat object and was then merged together. Dead cells and doublets were filtered by the following criteria: 1) cells with gene count less than 200 or more than 4000 gene were excluded; 2) cells with UMI count more than 20000 were excluded; 3) cells with mitochondrial content > 20% were excluded; 4) potential doublets were detected and removed by the Scrublet package with default parameters[17]. After filtering, 29930 cells were retained for the next analyses. Then Gene expression matrix was normalized with a scale factor 10,000 using Seurat “NormalizeData” functions. Subsequently, top 2000 variable genes were selected and a linear scaling method was applied to standardize the range of expression values for each gene. Principal component analysis (PCA) was used to reduce dimensionality by Seurat “RunPCA” function. For cells visualization, the Uniform manifold approximation and projection (UMAP) was performed on the top principal components (PCs) by Seurat “RunUMAP” function. Then, the batch effects from different platforms and donors were corrected by the Harmony (V1.0) package[18]. Based on the batch-corrected data, we used the Seurat “RunUMAP” function with max.dim = 1:30 and the Seurat “FindClusters” function with resolution 0.08 to generate different cell clusters.

### Major cell type annotation and subclustering

The following canonical markers were used for the annotation of major cell types: Keratinocytes: EPCAM, KRT5, KRT14; Melanocyte: PMEL, MELANA; T cells: CD3D, TRBC2; Myeloid cells: LYZ, CD68; B cells and plasma cells: JCHAIN, IGHA1, CD79A, MZB1; Mast cells: CPA3, IL1RL1; Neutrophil: CSF3R, FCGR3B; Fibroblasts: LUM, COL1A1; Pericytes/Smooth muscle cells: RGS5, ACTA2; Lymphatic endothelial cells: LYVE, PECAM1; Vessel endothelial cells: SELE, PECAM1. Further re-clustering was performed for keratinocytes, immune cells and fibroblasts for in-depth investigation and better annotation of clusters based on more specific markers within each cell type. The B/Plasma compartment was re-clustered based on the following markers: B cells: MS4A1, CD79A; Plasma cells: MZB1, IGHA1; Plasmacytoid DC: IL3RA, LILRA4 (Supplementary Fig. S2d-f). As proliferative cells, marked by TOP21, MKI67, tends to cluster various types of cells and thus lack of specificity, we didn’t put them into subsequent analysis. During the subclustering analysis, we inspected some potential doublets which expressed marker genes of two or more cell types and we didn’t put these potential doublets in subsequent analysis.

### Differentially expressed genes, GSEA and GO analysis

To identify the differentially expressed genes (DEGs) of the cell types, we compared the gene expression values of the cluster of interest to that of the rest of the clusters by the Seurat “FindMarkers” function. The marker genes of interest cell cluster were gained with the log2foldchange threshold at 0.25 and the genes were expressed in at least 25% of cells from the interest cluster. To describe the function features of cell subtypes, we gained the gene sets of 50 hallmarks from the MSigDB database[19] and performed gene set enrichment analysis (GSEA) on the results of DEGs analysis using the fgsea (V1.18.0) package[20]. For gene ontology (GO) analysis, we used the clusterProfiler (V4.1.4) package to evaluate pathway activities[21]. In both GSEA and GO analysis, p.adjust value of pathways < 0.05 was considered to be significantly enriched.

### Calculation of function module scores

For specific function modules, we calculated the scores for each single cell by the Seurat “AddModuleScore” function. We collected four gene sets including the cytotoxicity module, the exhausted/inhibitory module, the macrophage phenotype 1 (M1) module, and the macrophage phenotype 2 (M2) module. (Table S3)[22, 23]. To calculate the M1/M2 ratio, we used the Min-Max Normalization method to normalize the M1 and M2 scores. And we collected gene lists of GO terms from MSigDB[24] to evaluate the interested functions of T cells and macrophages including “B cell chemotaxis”, “Immune response” and so on. The statistical significance between sample types or cells was calculated by ggsinif package using Wilcoxon’s test. \**p* < 0.05, \*\**p* < 0.01, \*\*\**p* < 0.001.

### Cell-to-cell interactions

The CellChat (V1.1.3, https://github.com/sqjin/CellChat) algorithm was used to infer cell-cell interactions and identify differential interactions between tumor and adjacent normal samples[25]. In brief, we followed the official workflow and imported gene expression data of tumor and adjacent normal samples into CellChat using “createCellChat” function, separately. The “compareInteractions”, “netAnalysis_signalingRole_scatter”, “rankNet” functions were used to perform interaction comparison between tumor and adjacent normal samples and selected significant interactions. We mainly applied “netVisual_aggregate” function to show the specific interaction difference between the two sample types and applied the “plotGeneExpression” function to show the expression level of ligands and receptors genes. All cell interaction visualizations were plotted using the CellChat package.

### Immunohistochemistry staining

For immunohistochemical (IHC) staining, the samples collected from 39 patients with iBCC (Table S4) were sectioned for 4 um. Formalin-fixed paraffin-embedded (FFPE) sections were deparaffinized for 2 h 60 °C and re-hydrated, followed by epitope retrieval in citrate buffer (pH 6) in a pressure cooker. Antigen retrieval was facilitated by subsequent incubation in 150mM Tris-HCl at pH 9 for 15 minutes at 70°C. The sections were cooled to room temperature, washed with PBS, circled with an immunohistochemical pen (Zhongshan Jinqiao Biotechnology), and closed with peroxidase for 10 min. The peroxidase was discarded and the sections was washed with PBS, and sealed with goat serum (Zhongshan Jinqiao Biotechnology) for 1 h. Primary antibody, MDK antibody (Abcam), incubation was performed overnight at 4 °C in a humidified chamber. After washing, appropriate secondary antibodies and the AEC peroxidase substrate kit (Zhongshan Jinqiao Biotechnology) with DAB as a substrate (Zhongshan Jinqiao Biotechnology) were applied. Finally, the sections were dehydrated, cleared and mounted in aqueous mounting medium for microscopic evaluation.

### Semiquantitative analysis of MDK staining and measuring infiltration depth of iBCC

All of the samples were assessed by two independent pathologists with experience evaluating IHC who were blinded of the patients’ clinical outcomes. MDK expression was measured semiquantitatively by calculating the percentage of immunoreactive cells that stained positively and the staining intensity. The percentage of immunoreactive cells was rated as follows: 0 points, <10%; 1 point, 10–50%; 2 points, >50%. The staining intensity was rated as follows: 0 (no staining or weak staining = light yellow), 1 (moderate staining ¼ yellow brown) and 2 (strong staining = brown). The overall score for MDK expression was the sum of points determined for the percentage of positively stained immunoreactive cells and the expression, and an overall score ranging from 0 to 4 was assigned[26–28]. For the statistical analysis, the patients were divided into two groups: a low expression group and a high expression group. An overall score between 0 and 2 was defined as low expression and between 3 and 4 as high expression. The independent scores awarded by the two pathologists were merged into a final score, which was reported in this study. The infiltration depth was determined by measuring the vertical distance from the epithelium’s basal layer to the deepest point of tumor cell infiltration. Any disagreements in the evaluation were settled by discussion between the two pathologists.

### Statistical analysis

The correlation between MDK expression and infiltration depth of iBCC was analyzed by Spearman rank correlation analysis. The Fisher’s exact test was used to analyze the differences of clinic pathologic characteristics. The relationship between MDK expression and infiltration depth of iBCC was assessed by the OR and the 95% CI that were estimated using univariate and multivariate logistic regression with covariate adjustment. All statistical tests were two-tailed, and *p* < 0.05 were considered statistically significant. Statistical analyses in this study were performed using SPSS statistics 26 software.

## Results

### Single-cell transcriptome atlas of human iBCC and adjacent normal skin

To fully illustrate the cellular landscape of human BCC, we performed a scRNA-seq analysis of iBCC and ANS samples from five patients diagnosed with iBCC (Fig. 1a, Table S1). After quality control, filtering, and standard processing procedures (supplemental materials), we obtained the transcriptomic profiles of 29,334 cells (Fig. 1a, Table S2). Eleven major cell types were identified by canonical cell markers, including keratinocytes, melanocytes, T/NK cells, myeloid cells, B/Plasma cells, neutrophils, mast cells, fibroblasts, pericytes/smooth muscle cells, vascular endothelial cells (VECs), and LECs (Fig. 1b, c, supplemental materials). These abundant cell populations and various cell types allowed us to accurately elucidate the TME of iBCC. Generally, compared with ANS cells, myeloid cells and B/plasma cells showed elevated cell proportions, whereas fibroblasts displayed reduced cell abundance in iBCC cells (Fig. 1d, e). Subsequently, we focused on keratinocytes, lymphoid cells, myeloid cells and stromal cells, which are major components of the TME of iBCC, for the downstream analysis.

**Fig. 1.**
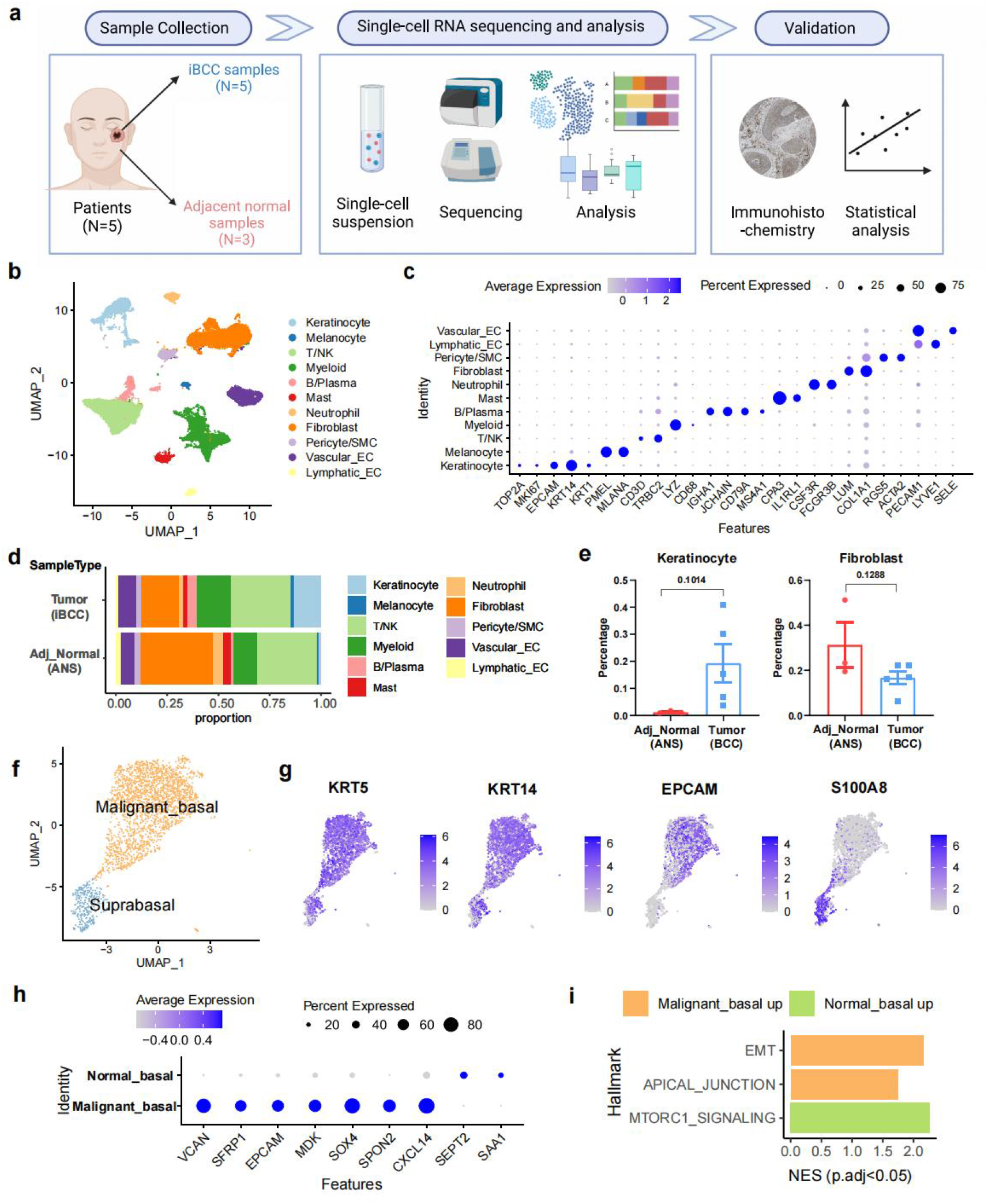
Single-cell atlas of human iBCC and ANS, and identification of malignant basal cells. (a) Graphical scheme describing the workflow. (b) The UMAP plot of major cell types in iBCC and ANS, different color represents different cell types. (c) Dot plot showing the expression levels of marker genes in each cell type. (d) The bar chart showed the proportion of cell types in ANS and iBCC. (e) The cell type population comparisons between two sample types. (f) The UMAP plot of keratinocytes sub-populations in iBCC. (g) The expression of marker genes of keratinocytes sub-populations in iBCC among UMAP. (h) Dot plot showing the differential expression genes of NBCs and MBCs. (i) Significantly enriched hallmark in malignant basal cells versus normal basal cells. Infiltrative basal cell carcinoma, iBCC; adjacent normal skin, ANS; EC, endothelial cell; NES, normalized enrichment score; NBCs, normal basal cells; MBCs, malignant basal cells.

### scRNA-Seq analyses recapitulate subpopulations of malignant basal cells and normal basal cells

As normal keratinocytes account for only a thin layer of the skin and are fairly eligible during tissue digestion, the number of keratinocytes obtained from ANS was far less than that from iBCC (33 cells vs. 2324 cells). To better define the keratinocyte group, we first analyzed them in iBCC and ANS separately. Using the basal cell markers KRT5 and KRT14, the suprabasal marker S100A8, and the classical tumor marker EPCAM, we identified normal basal cells (NBCs; KRT5^+^ and KRT14^+^) and normal suprabasal cells (S100A8^+^, KRT5^-^, and KRT14^-^) in the ANS (Supplementary Fig. S1e, f). Meanwhile, we identified MBCs (EPCAM^+^, KRT5^+^, and KRT14^+^) and suprabasal cells (S100A8^+^, KRT5^+^, and KRT14^+^) in iBCC (Fig. 1f, g). Notably, KRT5 and KRT14 were upregulated in the suprabasal cells of iBCC compared with those in ANS (Fig. 1g, Supplementary Fig. S1f). Next, we identified the differentially expressed genes and performed functional annotation to characterize the distinct functions of MBCs and NBCs. We found that MBCs expressed high levels of EPCAM, VCAN, SFRP1, MDK, SPON2, SOX4, and CXCL14 (Fig. 1h). In addition, the hallmark enrichment analysis showed that MBCs displayed significant “EMT” and “apical junction” signatures (Fig. 1i). Then, we calculated the genome-wide chromosomal copy number variations (CNVs) in NBCs and MBCs using inferCNV. We found that MBCs exhibited typical CNV alterations compared to NBCs (Supplementary Fig. 1g). Specifically, most MBCs showed depletion of CNV on chromosome 19, and the related genes were enriched in “oxidation phosphorylation” and “p53 binding” terms, emphasizing the tumor property of MBCs (Supplementary Fig. 1h).

### Enrichment of CXCL13^+^ Tfh-like cells and CD4^+^ Treg cells in iBCC

The tumor immune microenvironment (TIME) is sophisticated and multifefacial. To better understand the immune state in iBCC, we first performed a global analysis of T/NK cells in iBCC and ANS. First, we reclustered T/NK cells into five CD4^+^ T and three CD8^+^ T/NK subtypes by classical and specifically expressed gene markers (Fig. 2a, Supplementary Fig. S2a). Th five CD4^+^ T-cell clusters included (1) follicular helper-like T (CD4^+^CXCL13^+^ Tfh-like; CD4^+^, CXCL13^+^[29] and MAF^+^[30]), (2) T helper 1 (CD4^+^ Th1, characterized by the interferon response genes IFIT3, ISG15, and OASL), (3) regulatory T (CD4^+^ Treg; CD4^+^, FOXP3^+^, IL2RA^+^, and CTLA4^+^); (4) memory T (CD4^+^ Tmem; CD4^+^, IL7R^+^, and ANXA1^+^), and (5) MT1X^+^ memory T (MT1X^+^ CD4^+^ Tmem; CD4^+^, IL7R^+^, ANXA1^+^, and MT1X^+^). The three CD8^+^ T/NK-cell clusters included (1) CD8^+^ cytotoxic T (CD8^+^ Tc; CD8A^+^ and GZMA^+^), (2) FGFBP2^+^ NK (FGFBP2^+^ and GNLY^+^), and (3) XCL1^+^ NK (XCL1^+^ and GNLY^+^).

**Fig. 2.**
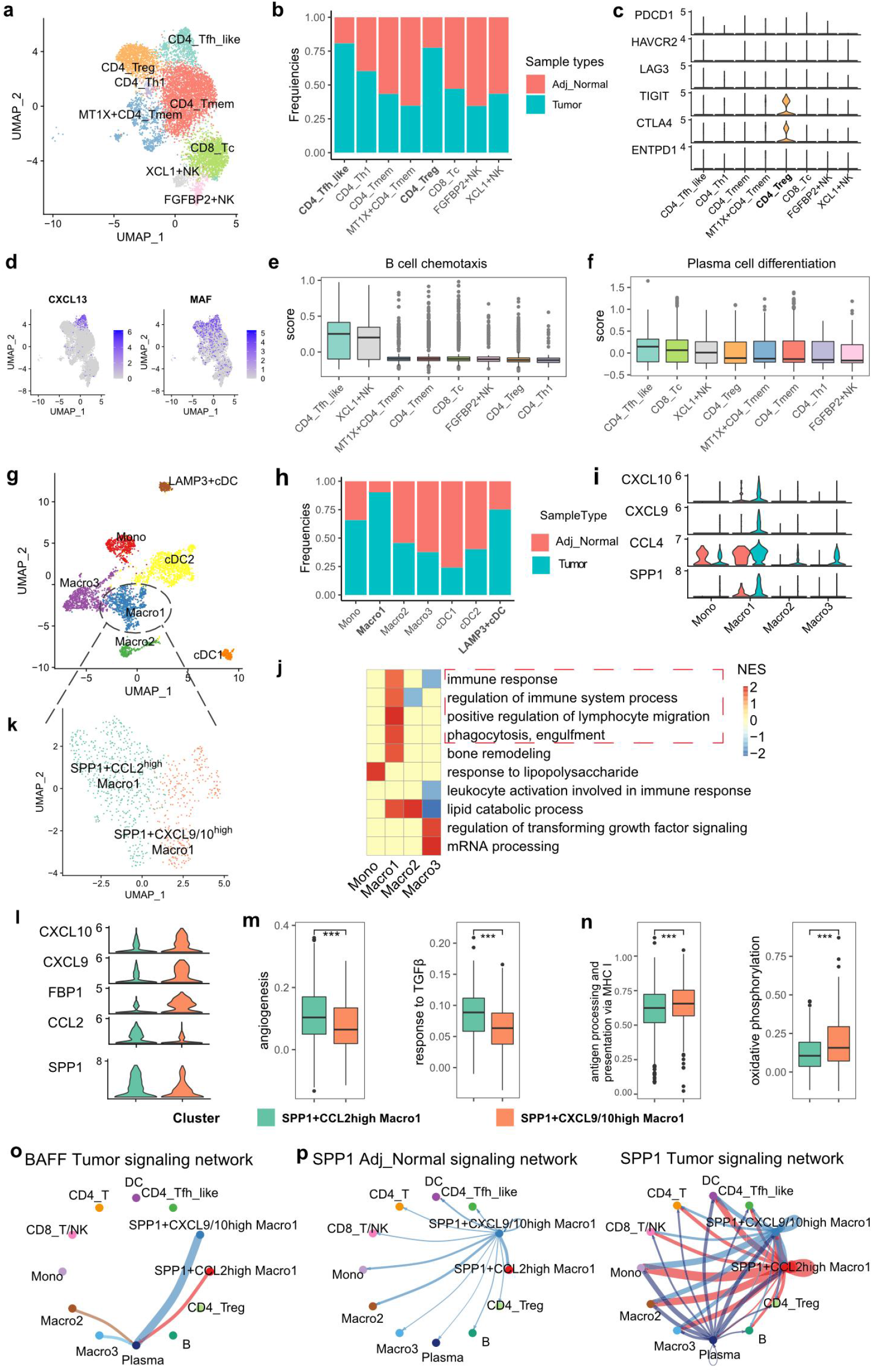
The immune landscape of iBCC at single-cell resolution. (a) UMAP visualization of T/NK cells. (b) Frequencies of cells from ANS and iBCC present in each cell subgroup in T/NK compartment (c) The expression level of immune inhibitors among different cell types. (d) The expression of marker genes of CD4^+^ Tfh-like cells among UMAP (e-f) The functional scores among different cell types (g) UMAP visualization of myeloid cells. (h) Frequencies of cells from ANS and iBCC present in each cell subgroup in myeloid compartment. (i) Violin plot showing the high expression marker genes of SPP1^+^ Macro1 between ANS and iBCC. (j) Heatmap of enrichment of GO terms in each macrophage subtypes. *p.adj* < 0.05. (k) UMAP visualization of two subtypes of SPP1^+^ Macro1. (l) High expression marker genes of two subtypes of SPP1^+^ Macro1. (m) Enriched GO term scores of SPP1^+^CCL2^high^ Macro1. (n) Enriched GO term scores of SPP1^+^CXCL9/10^high^ Macro1. (o) Circos plot showing the inferred cellular interaction network of BAFF signaling pathway in iBCC. (p) Cellular interaction network of SPP1 signaling pathway in ANS and iBCC. Wilcoxon signed-rank test, \**p* < 0.05, \*\**p* < 0.01, \*\*\**p* < 0.001. CD4^+^ Tfh, CD4^+^ follicular helper like T cell.

Notably, the CD4^+^ Tregs and CD4^+^CXCL13^+^ Tfh-like cells were enriched in iBCC compared to ANS (Fig. 2b). To characterize the immune activity of T/NK cells, we calculated the inhibitory/exhausted and cytotoxicity scores of each subtype (supplemental materials). CD4^+^ Treg cells had the highest inhibitory score among all T/NK subtypes, and the score was significantly upregulated in iBCC (*p* < 0.001, Supplementary Fig. S2b). Specifically, only CD4^+^ Treg cells exhibited a high expression of immune checkpoints, such as CTLA4 and TIGIT (Fig. 2c). CD4^+^CXCL13^+^ Tfh-like cells highly expressed CXCL13, a dominating chemoattractant in homing B cells to follicles[29]. Next, we evaluated the activity of the “B-cell chemotaxis” and “Plasma cell differentiation” pathways and found that the score of the CD4^+^CXCL13^+^ Tfh-like cells ranked first in both pathways among all T/NK subtypes, supporting its follicular helper cell identity. However, the inhibitory score of CD4^+^CXCL13^+^ Tfh-like cells was significantly increased in iBCC compared with that in ANS (*p* < 0.05), indicating an impaired function of CD4^+^CXCL13^+^ Tfh-like cells in the TME (Supplementary Fig. S2b).

Three CD8^+^ T/NK subtypes highly expressed cytotoxicity genes, such as NKG7 and GZMA, and chemoattractant genes, including CCL4 and CCL5. Nevertheless, these subtypes exhibited heterogeneous characteristics; FGFBP2^+^ NK cells and CD8^+^ Tc cells were the top two ranked subpopulations that possessed high cytotoxicity features (Supplementary Fig. S2c). The cytotoxicity score of CD8^+^ Tc and FGFBP2^+^ NK cells was reduced in iBCC cells compared with that in ANS cells (CD8^+^ Tc, *p* < 0.001; FGFBP2^+^ NK, *p* < 0.05, Supplementary Fig. S2c). Interestingly, CD8^+^ T/NK cells hardly expressed exhaustion markers and did not show a significant elevation in the exhaustion score in iBCC, indicating that the loss of toxicity in CD8^+^ Tc and FGFBP2^+^ NK cells was related to other mechanisms (Fig. 2c, Supplementary Fig. S2c). Taken together, among iBCC-enriched T cells, CD4^+^ Treg cells may be a key reason for the impaired immunity in iBCC; however, in contrast, CD4^+^CXCL13^+^ Tfh-like cells are potentially important factors in sustaining humoral immunity and maintaining the active immune function of iBCC.

### iBCC-enriched SPP1^+^ Macro1 displayed a proinflammatory role and showed heterogeneous phenotypes within the microenvironment

Recent single-cell profiling studies have revealed extensive heterogeneity in myeloid cells[22]. Across iBCC and ANS, myeloid cells were reclustered into three dendritic cells (DCs), one monocyte, and three macrophage subtypes (Fig. 2g). All Mono/Macro subtypes expressed CD14, whereas CD14 was almost absent in DCs. The monocyte subtype was characterized by a high expression of IL1B, S100A8, and EREG, and we distinguished macrophages from these cells by CD163 and MRC1 (Supplementary Fig. S3a). Three conventional DC subtypes were identified based on representative marker genes, including cDC1 (CLEC9A^+^ and S100B^+^), cDC2 (CLEC10A^+^, FCER1A^+^, and CD1C^+^) and LAMP3^+^ cDC (LAMP3^+^) (Supplementary Fig. S3a). Notably, LAMP3^+^ cDCs had activated and migratory markers, such as FSCN1, CCR7, and BIRC3, and harbored a higher cell abundance in iBCC cells than in ANS cells (Fig. 2h, Supplementary Fig. S3a).

Macrophages primarily function as phagocytic cells that can process and present antigens. Macrophages are highly plastic and have various phenotypes in the TME[22]. In this study, three subtypes of macrophages were detected according to the specific gene expression level and functions, including SPP1^+^ Macro1 (SPP1^+^, CCL4^+^, and CXCL9^+^), lipid-associated Macro2 (TGM2^+^, FABP4^+^, and LTA4H^+^), and FOLR2^+^ Macro3 (FOLR2^+^, F13A1^+^, and CCL13^+^) (Supplementary Fig. S3a).

Notably, SPP1^+^ Macro1 was obviously enriched in iBCC and had a high expression of CCL4, SPP1, CXCL9, and CXCL10 (Fig. 2h, i). Specifically, CXCL9 and CXCL10, as chemokines that recruit T/NK cells[31], were exclusively highly expressed in Macro1 cells in iBCC (Fig. 2i). Furthermore, by conducting a GO enrichment analysis, we found that Macro1 was enriched in inflammatory-associated pathways, including “immune response”, “positive regulation of lymphocyte migration”, and “phagocytosis, engulfment”. However, FOLR2^+^ Macro3 had high activities of “regulation of TGF signaling” but the lowest activities of “immune response” (Fig. 2j). SPP1^+^ macrophages were recently reported to be associated with angiogenesis, tumor metastases, and a worse prognosis[22, 32]. To characterize the heterogeneity of SPP1^+^ Macro1, we further reclustered Macro1 into two subgroups by their differentially expressed genes, namely, SPP1^+^CCL2^high^ Macro1 (SPP1^+^ and CCL2^high^) and SPP1^+^CXCL9/10^high^ Macro1 (SPP1^+^, CXCL9^high^, CXCL10^high^, and FBP1^high^) (Fig. 2k, l). Furthermore, we calculated the pathway scores of biological pathways of interest in the two subgroups to reveal their functions. Both subgroups had high scores in “inflammation response” and “phagocytosis” (Supplementary Fig. S3c). However, the SPP1^+^CCL2^high^ Macro1 group had significantly higher scores in “angiogenesis” and “response to TGF-β” (*p* < 0.001, Fig. 2m). SPP1^+^CXCL9/10^high^ Macro1 exhibited significantly higher activities of “antigen processing and presentation versus MHC-I” and “oxidative phosphorylation” (*p* < 0.001) (Fig. 2n). These results revealed that SPP1^+^ macrophages had heterogeneous functions in iBCC. Macrophages are traditionally divided into the proinflammatory type (M1) or the anti-inflammatory type (M2)[33, 34]. Then, we determined the M1/M2 scores of each macrophage subtype, and the results showed that SPP1^+^CXCL9/10^high^ Macro1 was the top-ranked subtype (supplemental materials), supporting their proinflammatory phenotypes. Overall, we characterized the myeloid atlas in iBCC and revealed the proinflammatory characteristic of SPP1^+^CXCL9/10^high^ Macro1 in the TME.

### SPP1^+^CXCL9/10^high^ Macro1 and SPP1^+^CCL2^high^ Macro1 displayed active communication with immune cells through different strengths of the BAFF and SPP1 signaling pathways

The TME comprises complex cellular interactions that include both activating and inhibiting signals. To further explore the effect of SPP1^+^CXCL9/10^high^ Macro1 and SPP1^+^CCL2^high^ Macro1 on other immune cells, we performed a cell-to-cell interaction analysis in iBCC and ANS. Generally, iBCC contains many more cell interaction events than ANS (Supplementary Fig. S3f). The incoming signaling strength of CD8^+^ T/NK, CD4^+^ Tfh-like, and CD4^+^ Treg cells was markedly increased in iBCC (Supplementary Fig. S3g). BAFF signaling and SPP1 signaling, which are intensively related to SPP1^+^CXCL9/10^high^ Macro1 and SPP1^+^CCL2^high^ Macro1, were significantly enriched in iBCC (Fig. 5b).

BAFF signaling, a critical pathway for B-cell activation and plasma cell survival[35, 36], was exclusively exhibited in iBCC (Fig. 2o). SPP1^+^CXCL9/10^high^ Macro1 had an obviously higher strength of BAFF signaling than SPP1^+^CCL2^high^ Macro1 (Fig. 2o). In detail, macrophage-derived TNFSF13B interacted with TNFRSF17 on plasma cells (Supplementary Fig. S3i).

SPP1^+^CCL2^high^ Macro1 showed relatively stronger SPP1 signaling than SPP1^+^CXCL9/10^high^ Macro1. The two subtypes of SPP1^+^ Macro1 displayed active communication with immune cells, especially myeloid cells, via the SPP1-CD44/integrin axis in iBCC (Fig. 2p, Supplementary Fig. S3j). In addition, plasma cells also expressed SPP1 in iBCC and, thus, exhibited extensive interactions with immune cells (Fig. 2p, Supplementary Fig. S3j). These results further reveal the heterogeneity of SPP1+ macrophages. Overall, we focused on SPP1^+^CXCL9/10^high^ Macro1 due to its strong immune-activation role and extensive interactions in the TIME.

### iBCC-associated ANGPT2^high^ lymphatic endothelial cells promoted leukocyte activation and migration

We extracted VECs, LECs, and pericytes/SMC compartments and reclustered them into the following six cell types based on classical markers: arterial ECs (SEMA3G^+^ and HEY1^+^), venous ECs (ACKR1^+^ and SELE^+^), capillary ECs (RGCC^+^), LECs (LYVE1^+^), pericytes (RGS5^+^), and SMCs (ACTA2^+^) (Fig. 3a, Supplementary Fig. S4a). Capillary ECs were more enriched in iBCC than in ANS (Fig. 3b).

**Fig. 3.**
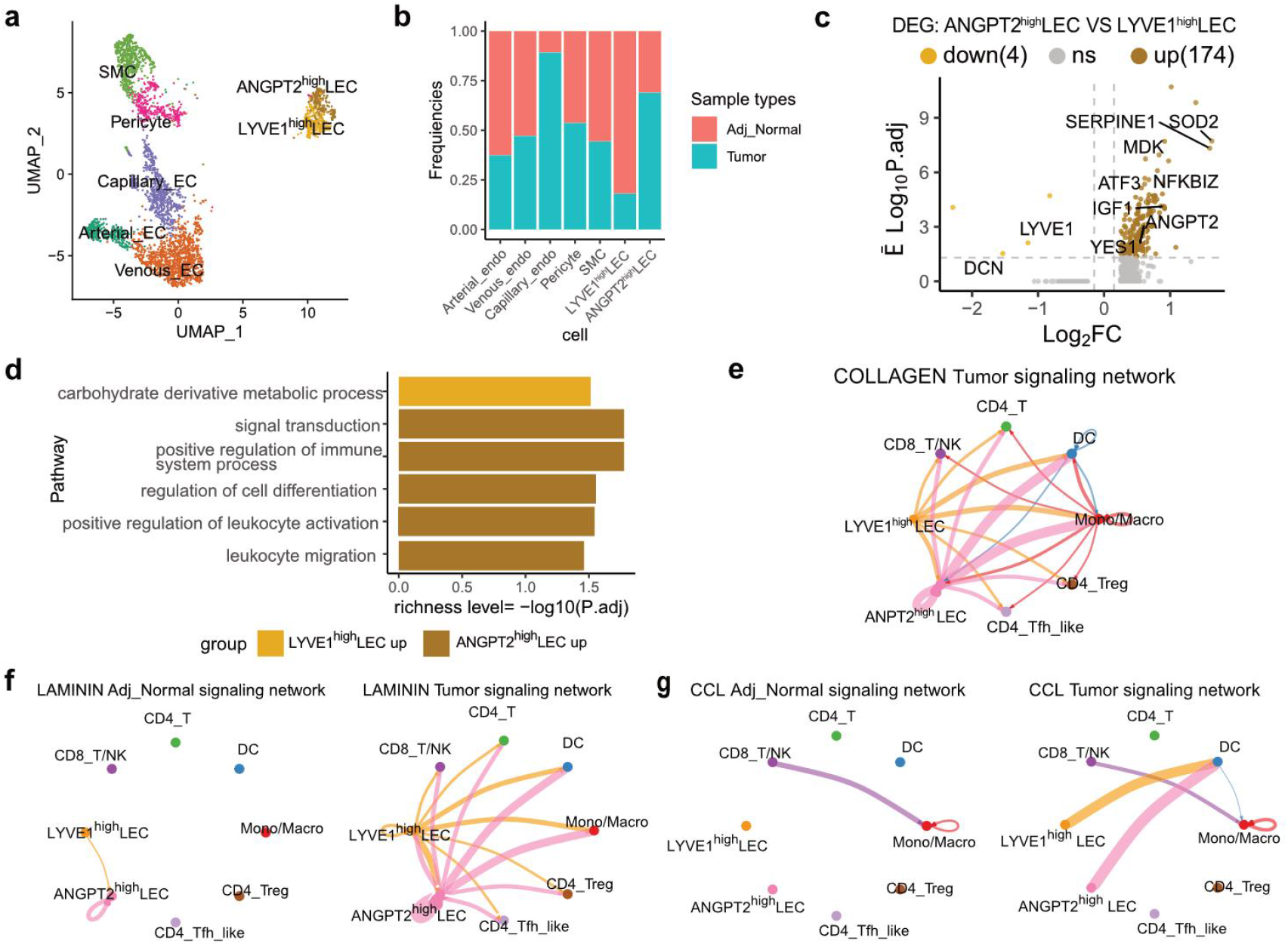
Tumor-associated ANGPT2^high^ lymphatic endothelial cells. (a) UMAP plot showing the subgroups of ECs, pericytes, and SMC. (b) Frequencies of cells from ANS and iBCC present in each cell subgroup. (c) Volcano plot showing the differential expressed genes between ANGPT2^high^ LEC and LYVE1^high^ LEC. (d) The enriched GO terms in LYVE1^high^ LEC versus ANGPT2^high^ LEC. (e) Cellular interactions of COLLAGEN signaling pathway in iBCC. (f) Cellular interactions of LAMININ signaling pathway in ANS and iBCC. (g) Cellular interactions of CCL signaling pathway in ANS and iBCC. LEC, lymphatic endothelial cell.

LECs were further classified into two clusters based on their distinct gene signatures. In addition to the classical LYVE1^high^ LECs, we newly identified ANGPT2^high^ LECs. ANGPT2^high^ LECs were enriched in iBCC and highly expressed inflammatory factors, such as SERPINE1, YES1, and NFKBIZ and the growth factor MDK (Fig. 3b, c). Additionally, the functional enrichment analysis revealed that ANGPT2^high^ LECs were enriched in the “leukocyte migration”, “positive regulation of leukocyte activation”, and “regulation of cell differentiation” pathways (Fig. 3d). To further explore the role of ANGPT2^-high^ LECs, we performed a cell communication analysis between LECs and immune cells in iBCC and ANS separately (Supplementary Fig. 3c). We observed three associated signaling pathways significantly enriched in iBCC, including COLLAGEN, LAMININ, and CCL21-CCR7 signaling (Supplementary Fig. S4b). Collagens and laminins are the major components of the extracellular matrix, and laminins compose the basal membrane upon endothelial and epithelial cells [37]. Collagens and laminins secreted from LECs bind CD44 on immune cells (T/NK cells and myeloid cells) and integrins. COLLAGEN signaling was iBCC specific, and ANGPT2^high^ LECs contributed more strongly than LYVE1^high^ LECs. The LAMININ signaling between ANGPT2^high^ LECs and immune cells was also iBCC specific. CCL21 in LECs recruited CCR7-expressing DCs, and this signaling was exclusive to iBCC (Fig. 3g, Supplementary Fig. S4f), which showed a slightly higher level in ANGPT2^-high^ LECs than in LYVE1^high^ LECs (Supplementary Fig. S4f). These observations indicate that ANGPT2^-high^ LECs play a more important role in interacting with immune cells. In conclusion, we identified a novel, tumor-associated ANGPT2^high^ LEC subtype that may play an important role in sustaining the high activities of immune cells and immune surveillance in iBCC.

### Diverse phenotypes and functions of fibroblasts in iBCC

In total, we detected 6299 fibroblast cells, which were further reclustered into six distinct subtypes by their gene signatures and functions, including (1) normal Fib1 (nFib; ADH1B^+^, APOD^+^, and PI16^+^), (2) transforming Fib2 (tFib; APCDD1^+^, F13A1^+^, and STC1^+^), (3) inflammatory cancer-associated Fib3 (iCAF; SERPINE2^+^, CCL2^+^, and CCL19^+^), (4) matrix-related cancer-associated Fib4 (mCAF; POSTN^+^, TNC^+^, and COL11A1^+^), (5) antigen-presenting Fib5 (apFib; CD74^+^, HLA-DRB1^+^, and HLA-DRA^+^), and (6) developing Fib6 (dFib; IGFBP3^+^ and WNT5A^+^) (Fig. 4a, d). Among the six subtypes, iCAFs and mCAFs were enriched in iBCCs, while nFibs were enriched in ANSs (Fig. 4b). The fibroblast subtypes, except for mCAFs, highly expressed the chemokines CXCL12 and CXCL14 (Fig. 4c). The “hallmark” enrichment analysis further supported our classification. tFib, iCAF, and apFib had high activities in inflammatory-associated pathways, including the “TNF signaling” and “complement” pathways (Fig. 4e). In addition, both iCAFs and mCAFs had high activities in the “EMT” and “angiogenesis” pathways (Fig. 4e).

**Fig. 4.**
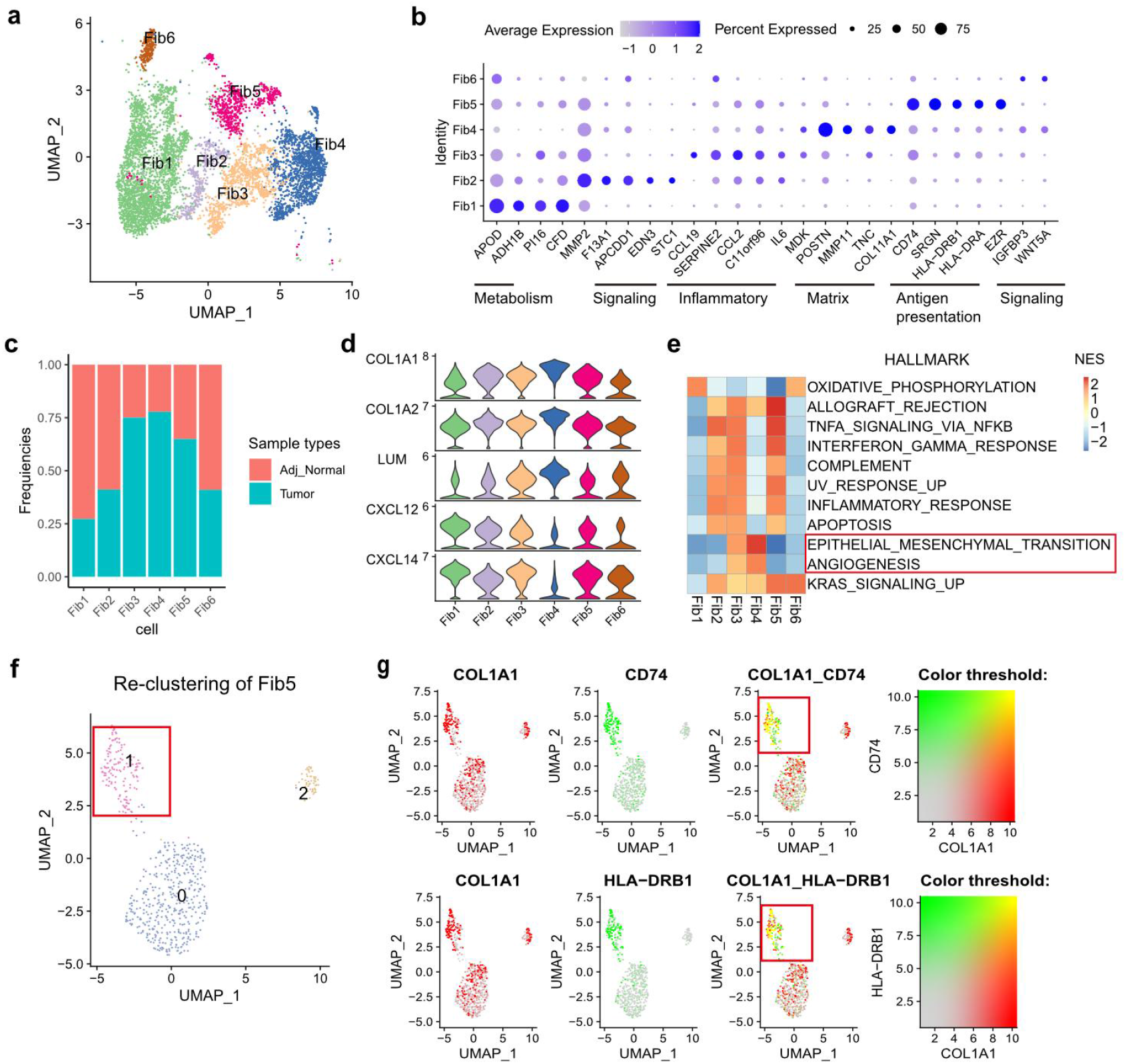
Diverse phenotypes of fibroblasts. (a) UMAP plot of fibroblast subgroups. (b) Differential expressed marker genes of fibroblast subgroups and corresponding functions. (c) Frequencies of cells from ANS and iBCC present in each cell subgroup. (d) Volin plot showing the commonly expressed genes in fibroblasts subgroups. (e) Heatmap showing the enriched hallmark scores of each fibroblasts subgroup. *P.adj* < 0.05. (f) UMAP visualization of sub-cluster 0, 1, 2 from Fib5. (g) The split and merged expression level of two genes in Fib5 among UMAP. The colorization of yellow on cell dots signifying the co-expression level of the two genes. Fib, fibroblast.

Antigen-presenting fibroblasts were recently identified in malignancies and adjacent normal tissues and could activate CD4^+^ T cells[38–40]. Here, we found that apFib expressed both fibroblast (LUM, COL1A1, and COL1A2) and antigen presentation-associated genes (CD74, HLA-DRB1, and HLA-DRA) (Fig. 4d). Meanwhile, apFib had similar gene and UMI numbers compared to the other subtypes, which could exclude their doublet identity (Supplementary Fig. S5a). We further extracted apFib and reclustered them into subclusters 0, 1, and 2 (Fig. 4f). Subcluster 1 of apFib had the highest coexpression level of COL1A1 and CD74/HLA-DRB1 (Fig. 4g), while subcluster 0 of apFib highly coexpressed COL1A1 and PTPRC/CXCR4 (Supplementary Fig. S5b). Taken together, the above results provide an integrated picture of diverse fibroblasts across iBCC and ANS.

### Globally cell-to-cell interaction analysis revealed antigen presentation-associated fibroblasts in iBCC

Next, we performed a global cellular interaction analysis of major cells in iBCC and ANS and detected differential interaction relationships. Generally, the cellular interaction network in iBCC was much more complex than that in ANS (Fig. 5a). Four signaling pathways were significantly upregulated in iBCC, including CD99, LAMININ, MHC-I, and MDK signaling (Fig. 5b). Fibroblasts, the major transmitters of the chemokine CXCL12, recruited various CXCR4-expressing immune cells into tissues in iBCC and ANS (Supplementary Fig. S6a, b). CD99 signaling involves the CD99-CD99 and CD99-PILRA ligand–receptor pairs and is crucial for the transmigration of leukocytes through the vessel wall[41]. CD99 signaling was widely enhanced in iBCC (Fig. 5c). Specifically, CD4^+^CXCL13^+^ Tfh-like cells and B/plasma cells displayed an exclusively high expression pattern of CD99 in iBCC, while the expression of CD99 was downregulated in MBCs and CD4^+^ Tregs in iBCC (Fig. 5d). LAMININ signaling displayed an iBCC-specific interaction pattern in ECs and MBCs and was enhanced in fibroblasts in iBCC versus ANS (Fig. 5e, Supplementary Fig. S6d). The difference in CD99 and LAMININ signaling between iBCC and ANS showed a complicated regulatory network of cell adhesion.

**Fig. 5.**
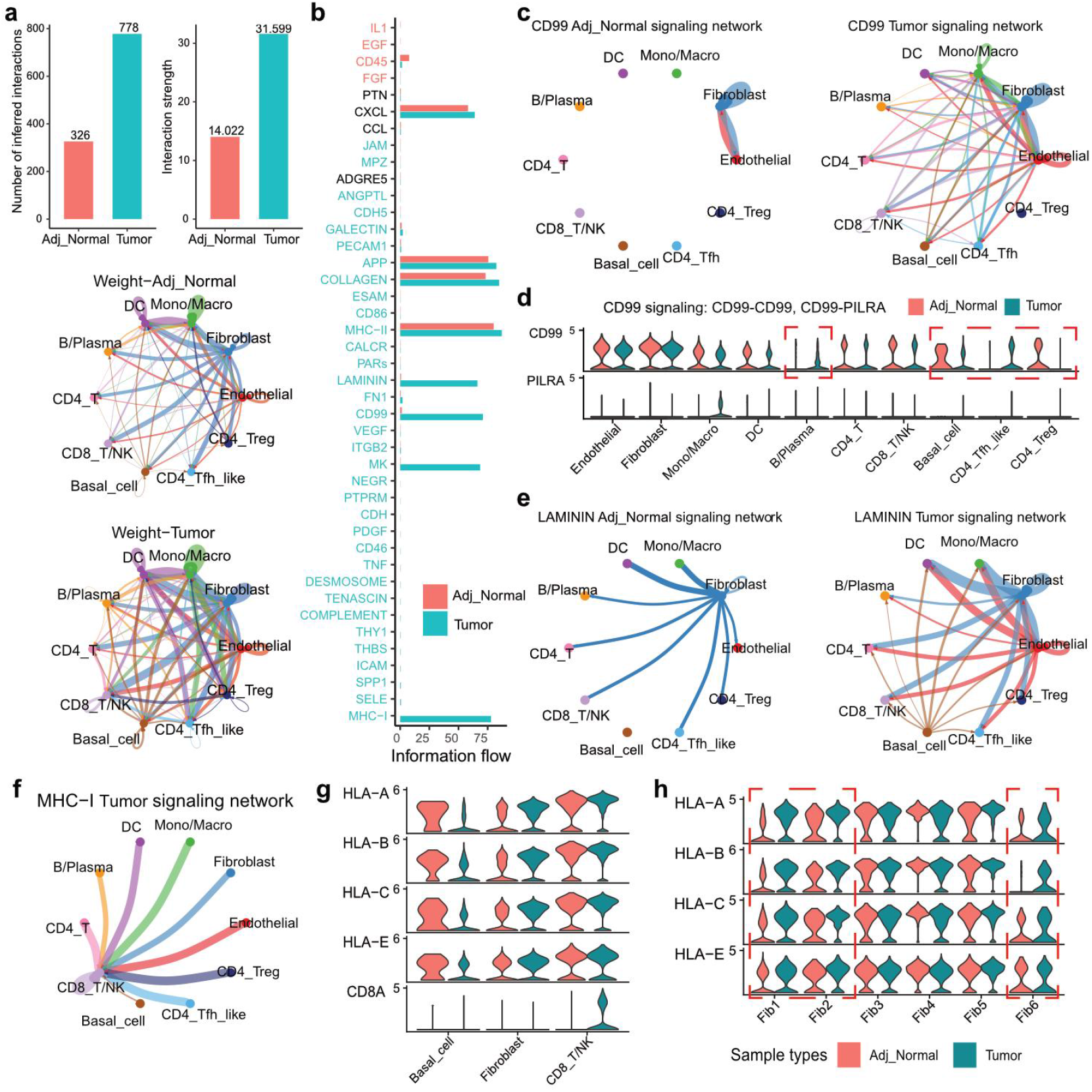
Globally cellular interactions in iBCC and ANS. (a) The number, strength, network of inferred cellular interactions among major cell types in ANS and iBCC. (b) The significant signaling pathways were ranked based on their differences in overall information flow within the inferred networks between ANS and iBCC. (c) The circus plot showing the cellular interactions of CD99 signaling pathway. (d) The expression of ligands and receptors among different cell types of CD99 signaling, and colors represent different cell types. (e) Cellular interactions of LAMININ signaling pathway in ANS and iBCC. (f) Cellular interactions of MHC-I signaling pathway. (g) The expression of ligands and receptors of MHC-I signaling, and colors represent different sample types. (h) The expression of MHC-I genes among fibroblast subgroups from different sample types.

The MHC-I signaling (HLA-CD8A)-related interaction was iBCC specific, its ligand was widely expressed in each cell, and the receptor CD8A was exclusively expressed in CD8^+^ T/NK cells, resulting in a strong interaction between CD8^+^ T/NK cells and other cells and an intercellular relationship within CD8^+^ T/NK cells (Fig. 5f). Interestingly, we found that MHC-I genes (such as HLA-A and HLA-B) were downregulated in MBCs, whereas fibroblasts harbored an elevated expression level of MHC-I genes in iBCC compared with that in ANS (Fig. 5g). In detail, nFib (Fib1) and tFib (Fib2) had a significantly higher expression of MHC-I genes in iBCC versus ANS (Fig. 5h). MBCs express low levels of MHC-I genes and, thus, escape CD8+ T/NK killing. However, fibroblasts highly expressed MHC-1 genes, potentially increasing the activity of CD8+ T cells, exerting immune surveillance and attenuating tumor cell escape, thus maintaining a highly activated immune microenvironment.

### Tumor-derived MDK remodeled the tumor microenvironment and was positively correlated with iBCC infiltration

MBCs established strong connections with themselves and other cells in iBCC by MDK signaling, but their interaction strength was weak in the ANS (Fig. 6a). Among them, the signaling strength between MBCs and fibroblasts was the strongest. MDK, a secreted growth factor, can bind various receptors and promote tumor growth during tumorigenesis[42–45] (Fig. 6b). MBCs autocrine MDK, binding SDC receptor on themselves and fibroblasts. Fibroblasts, macrophages and endothelial cells could also express other receptors to communicate with MBCs (Fig. 6b).

**Fig. 6.**
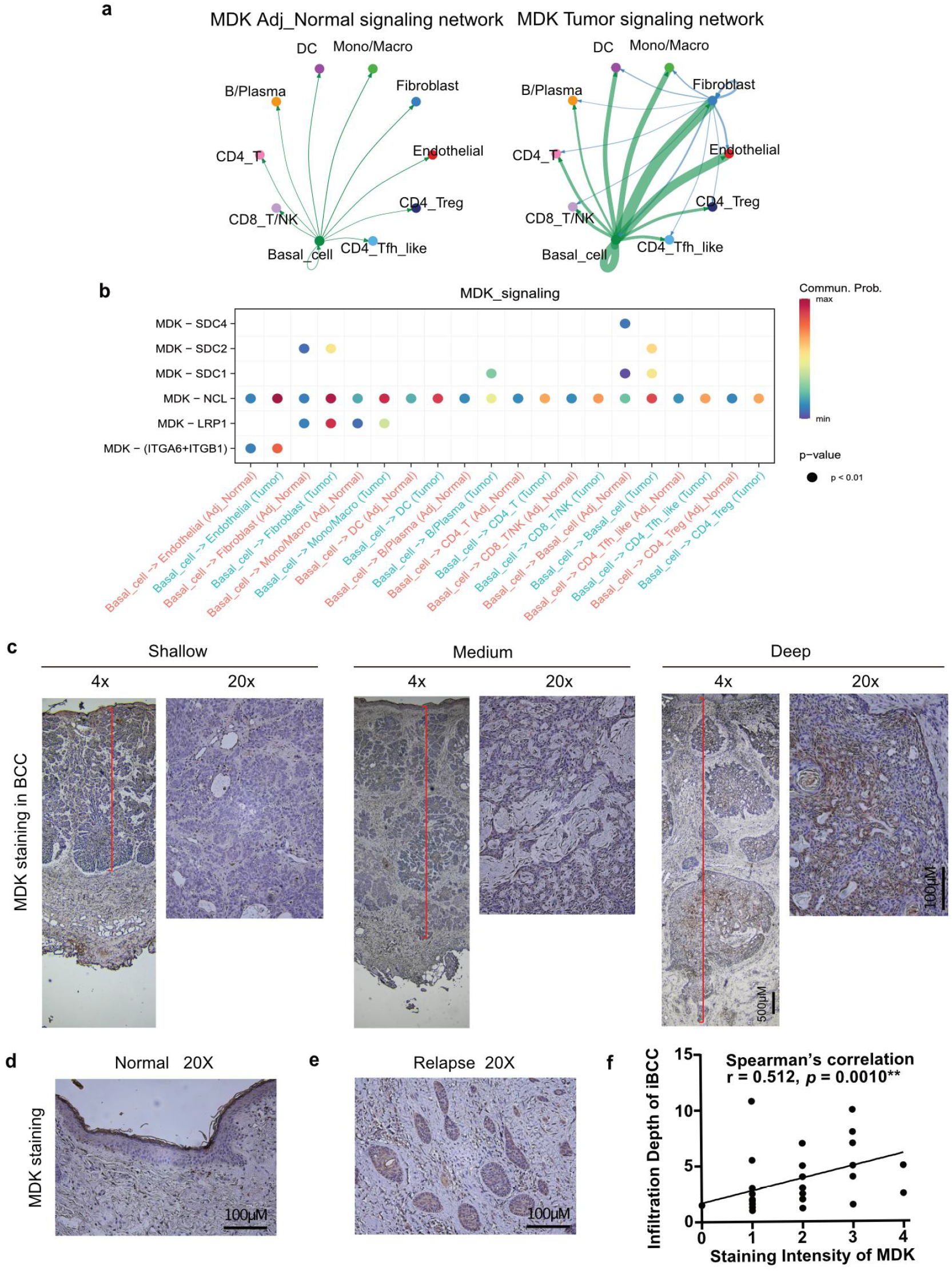
Tumor-derived MDK influenced the TME. (a) Cellular interactions of MK signaling pathway in ANS and iBCC. (b) Ligand-receptor interaction probabilities of MK signaling. (c) IHC of MDK in defferent infiltration depth of iBCC. (d) IHC of MDK in normal skin. (e) IHC of MDK in reccurent iBCC. (f) Spearman’s rank correlation analysis between MDK staining intensity and iBCC infiltration depth. \*\**p* < 0.01.

To further explore the role of MDK, we performed IHC staining of MDK and measured the infiltration depth of tumors in a clinical cohort comprising 39 patients diagnosed with iBCC (supplemental materials). In addition, the clinical information of these patients was collected (Table S4, S5). We found that iBCC with different infiltration depths showed different expression levels of MDK, whereas MDK was barely expressed in NBCs (Fig. 6c, d). The expression level of MDK was significantly positively correlated with the infiltration depth of iBCC (r = 0.512, *p* < 0.01, Fig. 6f). Moreover, we identified MDK as an effective factor in predicting the risk of the infiltration depth of iBCC (odds ratio = 13.59, 95% confidence interval, 1.832, 100.785), which is independent of sex, age, duration of the tumor, tumor ulcer, and recurrence (Table 1). Overall, our results demonstrate that MDK is an independent risk factor for predicting the depth of iBCC invasion and is a key factor in tumor growth.

**Table 1.** Multivariate analysis of the association between the infiltration depth of iBCC and MDK expression.

| Variables | | B | SE (B) | $\chi^2$ | *p* | OR (95%CI) |
| --- | --- | --- | --- | --- | --- | --- |
| MDK expression | Low |  |  |  |  | 13.59 (1.832, 100.785) |
|  | High | 2.609 | 1.022 | 6.515 | 0.011 |  |
| Ulcer | No |  |  |  |  | 0.802 (0.078, 8.262) |
|  | Yes | -0.22 | 1.19 | 0.034 | 0.853 |  |
| Gender | Female |  |  |  |  | 1.307 (0.216, 7.913) |
|  | Male | 0.268 | 0.919 | 0.085 | 0.771 |  |
| Age (years) | <60 |  |  |  |  | 0.991 (0.908, 1.081) |
|  | ≥60 | -0.76 | 1.043 | 0.531 | 0.466 |  |
| Duration (years) | <5 |  |  |  |  | 1.415 (0.187, 10.701) |
|  | ≥5 | 0.347 | 1.032 | 0.113 | 0.736 |  |
| Reccurence | No |  |  |  |  | 6.328 (0.613, 65.298) |
|  | Yes | 1.845 | 1.191 | 2.401 | 0.121 |  |

The infiltraion depth division: < 5mm, ≥ 5 mm; Binary Logistic Regression; B and SE(B),the parameter estimator of association coefficient and its standard error; $\chi^2$ , Chi-squarestatistic; OR,odds ratio; CI, confidence interval

### Specific hallmarks and spatial gene expression distribution of the two heterogeneous subtypes of malignant basal cells

MBCs were further reclustered into two subtypes by specifically expressed genes and phenotypes, namely, SOSTDC1+IGFBP5+CTSV+ MB1 and TNC+SFRP1+CHGA+ MB2 (Fig. 7a, b). All iBCC samples contained these two subtypes but with different proportions, showing intratumor heterogeneity (Fig. 7c). By conducting a functional annotation analysis, we found that MB1 was enriched in the “keratinocyte differentiation” and “programmed cell death” pathways, while MB2 was enriched in the “extracellular matrix” and “skeletal system development” pathways (Supplementary Fig. S7a). Furthermore, we characterized MB1- and MB2-specific gene expression signatures and hallmarks using NBCs as a control. MB1 specifically expressed SOSTDC1, IGFBP5, CTSV, DSG1, and GADD45G, while MB2 displayed high expression levels of SFRP1, CHGA, COL14A1, and TNC (Fig. 7d, e). Moreover, MB1 had high activities of the “interferon response”, “TNF signaling”, “UV response up”, and “mTORc” pathways, which is similar to NB cells. MB2 had high activities of “EMT”, “mitotic spindle”, “apical junction”, and “angiogenesis pathways”, indicating its obvious malignant feature (Fig. 7f).

**Fig. 7.**
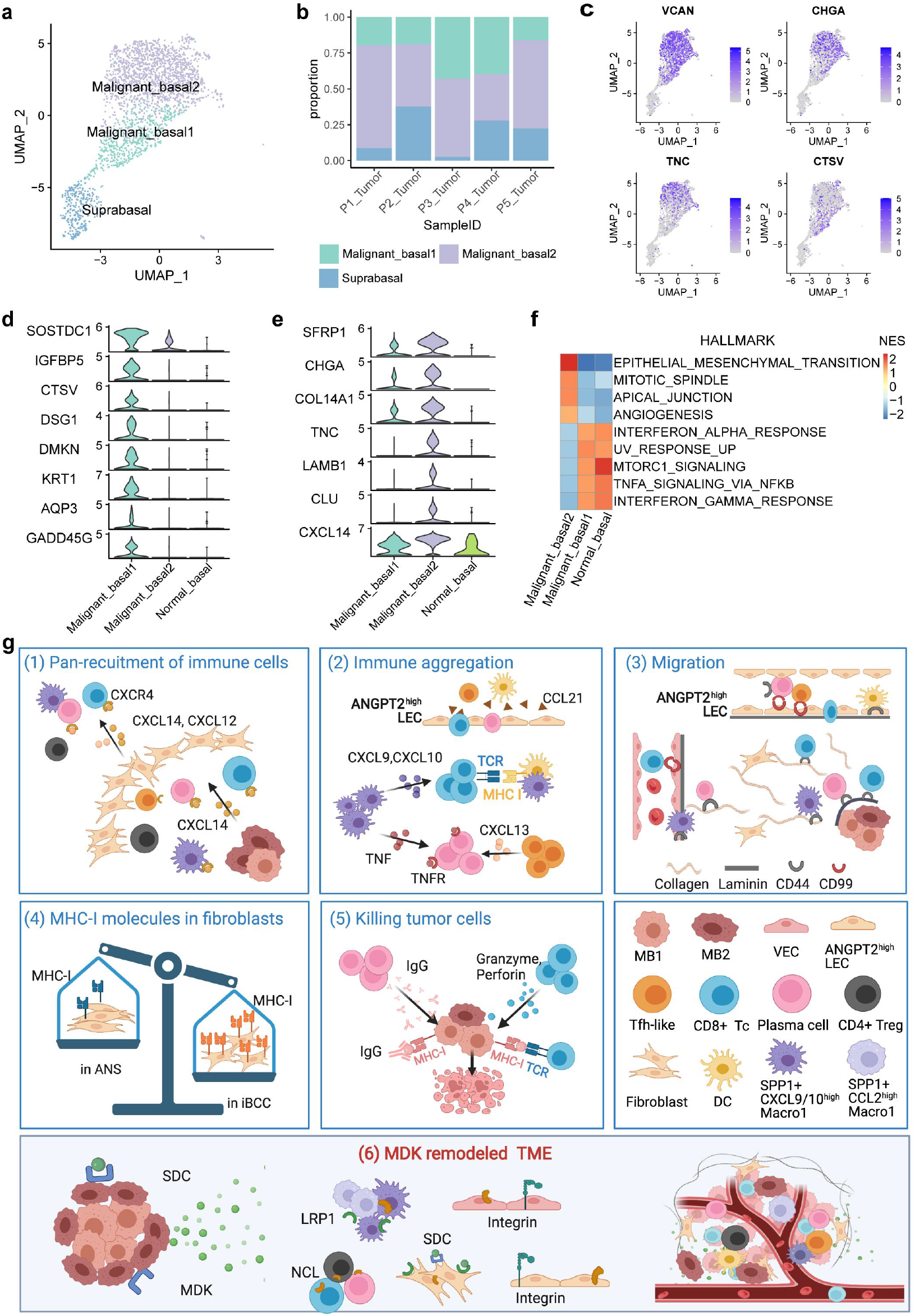
Two subtypes of malignant basal cells. (a) The UMAP plot of keratinocytes sub-populations in iBCC. (b) The proportion of keratinocytes sub-populations in each iBCC sample. (c) The expression of gene markers of MB1 and MB2 among UMAP. (d) The expression of gene markers of MB1 with NBCs as controls on Violin plot. (e) The expression of gene markers of MB2 with NBCs as controls on Violin plot. (f) Heatmap showing the enriched hallmark scores of MB1, MB2, and NBCs. *P.adj* < 0.05. g Schematic diagram of the major cell phenotypes and cell-cell interactions in iBCC. MB1, malignant basal cell type 1. MB2, malignant basal cell type 2.

To validate these gene signatures, we retrieved the IHC staining results from the Human Protein Atlas (https://www.proteinatlas.org/). Surprisingly, we found that the gene expression had a different spatial distribution. VCAN, a marker of MBCs, was highly expressed in the whole tumor (Fig. 7b, Supplementary Fig. S7a). CTSV, a marker of MB1, was mainly expressed in the tumor interior, whereas CHGA and TNC, markers of MB2, were mainly present in the peripheral tumor (Fig. 7c, d, Supplementary Fig. S7c, d, e). The IHC results could be explained by our scRNA-seq analysis. MB1 had a strong ability of epithelial differentiation and, therefore, tended to grow upward, thus being wrapped in the interior. In contrast, MB2 tends to lose cellular connections, have stronger motility, and undergo the epithelial mesenchyme transition; thus, MB2 moves to the peripheral area of the tumor.

## Discussion

In this study, we performed a comprehensive scRNA-seq analysis of iBCC and ANS and mainly depicted the dynamic cell heterogeneity and interactions among immune cells, myeloid cells, endothelial cells, and stromal cells. We revealed the inflammatory characteristics of the TME in iBCC, which was enriched with CD4^+^CXCL13^+^ Tfh-like cells, SPP1^+^CXCL9/10^high^ Macro1, SPP1^+^CCL2^high^ Macro1, and ANGPT2^+^ LECs. For the first time, we discovered that iBCC contained a subtype of antigen presentation-associated fibroblasts that potentially recruit and activate immune cells. More importantly, we revealed that malignancy-associated MDK signaling promotes iBCC infiltration and TME reprogramming. Two molecular types of iBCC with different malignant levels were also identified. The MB1/MB2 ratio might be an effective predictor of the malignancy level and prognosis of iBCC.

T-cell exhaustion has been demonstrated in a wide variety of human cancers and is characterized by the loss of effector and cytotoxic functions[46, 47]. A previous study found that both CD4^+^ and CD8^+^ T cells exhibited high inhibitory or exhausted states in advanced BCC[48]. However, in iBCC, CD4^+^ T cells showed a prominent inhibitory state, and CD8^+^ T cells did not show an obvious exhausted state. This evidence suggests that distinct T-cell states are involved in BCC at different stages, and their transforming characteristics may be related to the progression of BCC, ranging from the early stage of iBCC to advanced BCC.

Our study revealed active immune collaborations within iBCC. Specifically, compared with NBCs, MBCs had a high expression of CXCL14, a newly discovered ligand for CXCR4[49]. Meanwhile, all fibroblasts, except for mCAFs, showed a high expression of CXCL12 and CXCL14, which are two valid ligands for CXCR4 that are expressed on the surface of immune cells. Thus, the two chemokines may act as important regulators attracting immune cell infiltrates into the mesenchyme and further around MBCs. Interestingly, there were multiple active interactions among iBCC-enriched immune cells; for instance, CD4+CXCL13+ Tfh-like cells could recruit plasma cells, and SPP1^+^CXCL9/10^high^ Macro1 sustained the activities of plasma cells through BAFF signaling. In addition, SPP1^+^CXCL9/10^high^ Macro1 released CXCL9/10 to attract T/NK cells and further activate them through MHC-I signaling. CXCL13 [35, 50–52], CXCL9, CXCL10[53], and BAFF[35] have been shown to be associated with the formation of tertiary lymphatic structures (TLSs). TLS refers to organized aggregates of immune cells that form postnatally in nonlymphoid tissues, which often experience chronic inflammation and antigen persistence, and more TLSs are associated with a favorable prognosis in cancer[53]. Our results revealed that iBCC exhibited a “hot” tumor and displayed active immune cell collaboration within the TME, which may be harbored within immune aggregates (Fig. 7 Section 1, 2).

Recent studies reported that SPP1^+^ macrophages were associated with angiogenesis [22], tumor metastases [32], and worse survival in colorectal cancer (CRC)[54] and glioblastoma[55]. Unexpectedly, two subtypes of SPP1^+^ Macro1 in our study presented proinflammatory phenotypes, including the strongest immune chemotactic and phagocytic functions. SPP1^+^CCL2^high^ Macro1 had a higher angiogenesis capacity and response to TGF-β, while SPP1^+^CXCL9/10^high^ Macro1 had a stronger antigen presentation via MHC-I capability. Notably, both subtypes were almost absent in the ANS, demonstrating that they might be newly differentiated subtypes in iBCC. Macrophages are tremendously plastic and can differentiate into multiple phenotypes in different environmental contexts[56]. Therefore, we conceived that SPP1^+^ macrophages may differentiate in distinct directions and that their final phenotype depends on the specific TME. For example, in low-malignancy iBCC, the two subtypes of SPP1^+^ macrophages played a proinflammatory role. However, in CRC, the angiogenesis subtype assumes a prominent place, and thus, SPP1^+^ macrophages exhibit a protumor phenotype. In addition, previous studies found that SPP1 was associated with a better immunotherapy response in lung cancer[57] and a favorable prognosis in cholangiocarcinoma[58]. In non-small-cell lung cancer, researchers defined an activation module consisting of PDCD1^+^ CXCL13-activated T cells, IgG plasma cells, and SPP1^+^ macrophages, and a high baseline of this module was related to a better immune response[57]. On the basis of previous studies, we revealed the proinflammatory function of SPP1^+^CXCL9/10^high^ macrophages in low-malignancy iBCC.

Notably, we identified a novel tumor-associated ANGPT2^high^ LEC subtype that could promote the migration and activation of leukocytes. Compared to classical LYVE1^high^ LECs, ANGPT2^high^ LECs expressed more collagen-associated and laminin-associated genes. These components support the formation of lymphatic vessels[37] and promote the adhesion and migration of immune cells[59]. Therefore, ANGPT2^high^ LECs may play an important role in transporting immune cells to tumor tissues and exerting antitumor effects (Fig. 7, Section 2). Interestingly, ECs, MBCs, and fibroblasts showed elevated laminin signaling in iBCC, and CD99 signaling was widely enhanced in iBCC. CD99 signaling can facilitate the transmigration of immune cells through endothelial cells[41]. In iBCC, the expression of CD99 on B/plasma and CD4^+^ Tfh-like cells was upregulated but showed reduced expression in MBCs and CD4^+^ Treg cells, indicating that CD99 serves as an important adhesion molecule and is plastic in the TME. Taken together, we built a cell transmigration model in iBCC. Immune cells transmigrate ECs into tissues by LAMININ and CD99 signaling, move along the cellular skeleton built by fibroblasts, and finally land on laminins under MBCs, exerting a collaborative antitumor function (Fig. 7, Section 3).

For the first time, we discovered a subpopulation of antigen presentation-associated fibroblasts in the TME of iBCC that expressed high levels of MHC-I molecules. MHC-I molecules are widely present in a variety of cells, and DCs can cross-present antigens to CD8^+^ T cells and activate them via MHC-I molecules. It was newly reported that hepatic stellate cells could deliver MHC-I molecules with target antigens to hepatic sinusoidal endothelial cells, which, in turn, activated CD8^+^ T cells[50]. Multiple subtypes of fibroblasts with different phenotypes have been identified at the single-cell level, including antigen-presenting fibroblasts that carry high levels of MHC-2 molecules[38–40]. Whether the upregulated MHC-I molecules in fibroblasts play a role in activating CD8^+^ T/NK cells and the associated activation mechanisms need to be further explored (Fig. 7, Section 4). Taken together, the immune aggregates, cell migration, and potential antigen-presenting role of fibroblasts effectively play an immune surveillance role in iBCC (Fig. 7 Section 5).

MDK could induce macrophages to differentiate into an anti-inflammatory phenotype [45, 60] and promote the EMT[61] of cancer cells and angiogenesis[62, 63], which are associated with worse survival in many malignancies[45, 60]. Astonishingly, we found that the malignancy-associated MDK signaling pathway was upregulated in iBCC, and we further defined MDK expression as an independent risk factor for iBCC invasion, revealing MDK as a crucial malignancy driver of iBCC. To optimize treatment for BCC, European consensus-based interdisciplinary guidelines proposed a classification model dividing BCC into ‘easy-to-treat’ and ‘difficult-to-treat’ subtypes[2]. The ‘easy-to-treat’ BCC subtype refers to the most common BCC, and the ‘difficult-to-treat’ BCC subtype includes locally advanced (unresectable) BCC and common BCC posing specific management problems, such as those located in organs whose functions are easily damaged or BCCs with multiple recurrences. Therefore, ‘difficult-to-treat’ BCCs are more concerning. iBCC tends to progressively invade tissue and develop into locally advanced BCC, which has a higher risk of recurrence. Thus, elucidating the heterogeneity of iBCC is especially important. The identification of differentiation-associated SOSTDC1^+^IGFBP5^+^CTSV^+^ MB1 and EMT-associated TNC^+^SFRP1^+^CHGA^+^ MB2 further provides the molecular signature classification of iBCC with different degrees of malignancy. MB1 and MB2 had different proportions among the iBCC samples, and we believe that the MB1/MB2 ratio may be beneficial for the established treatment system and serve as a predictor of the progression and recurrence of iBCC (Fig. 7, Section 6).

## Conclusions

Overall, our study comprehensively depicted the diverse cellular phenotypes and cell-to-cell interactions in iBCC at a single-cell resolution, providing a fundamental reference for further research investigating the clinical issues of iBCC.

## Data Availability

The IHC results generated during this study are provided as supplementary tables. single-cell transcriptome and additional datasets are available upon request.

## Acknowledgements

We would like to thank the patients and their families for consenting tumor acquisition in our study.

## Contributions

Mingzhu Yin and Xiang Chen conceived and supervised this project. Lingjuan Huang, Xianggui Wang and Xin Li analyzed the scRNA-seq data. Xianggui Wang collected clinical samples and supervised the experiment. Shiyao Pei prepared the single-cell suspensions and did IHC staining. Lingjuan Huang, Mingzhu Yin, Xiang Chen, Xianggui Wang, Shiyao Pei and Xin Li wrote the manuscript. Liang Dong, Xiaohui Bian, Hongyin Sun, Liang Dong, Liping Jin, Huihui Hou organized figures and supervised the experiment. Wensheng Shi, Xiyuan Zhang, Lining Zhang and Shuang Zhao edited the manuscript.

## Ethics declarations

### Ethics approval and consent to participate

All patients were from Xiangya Hospital of Central South University and signed an informed consent form for providing samples. Our study was approved by the institutional ethics committe of Xiangya Hospital of Central South University (2022020109).

### Consent for publication

Not applicable.

---

**Fig. S1.**
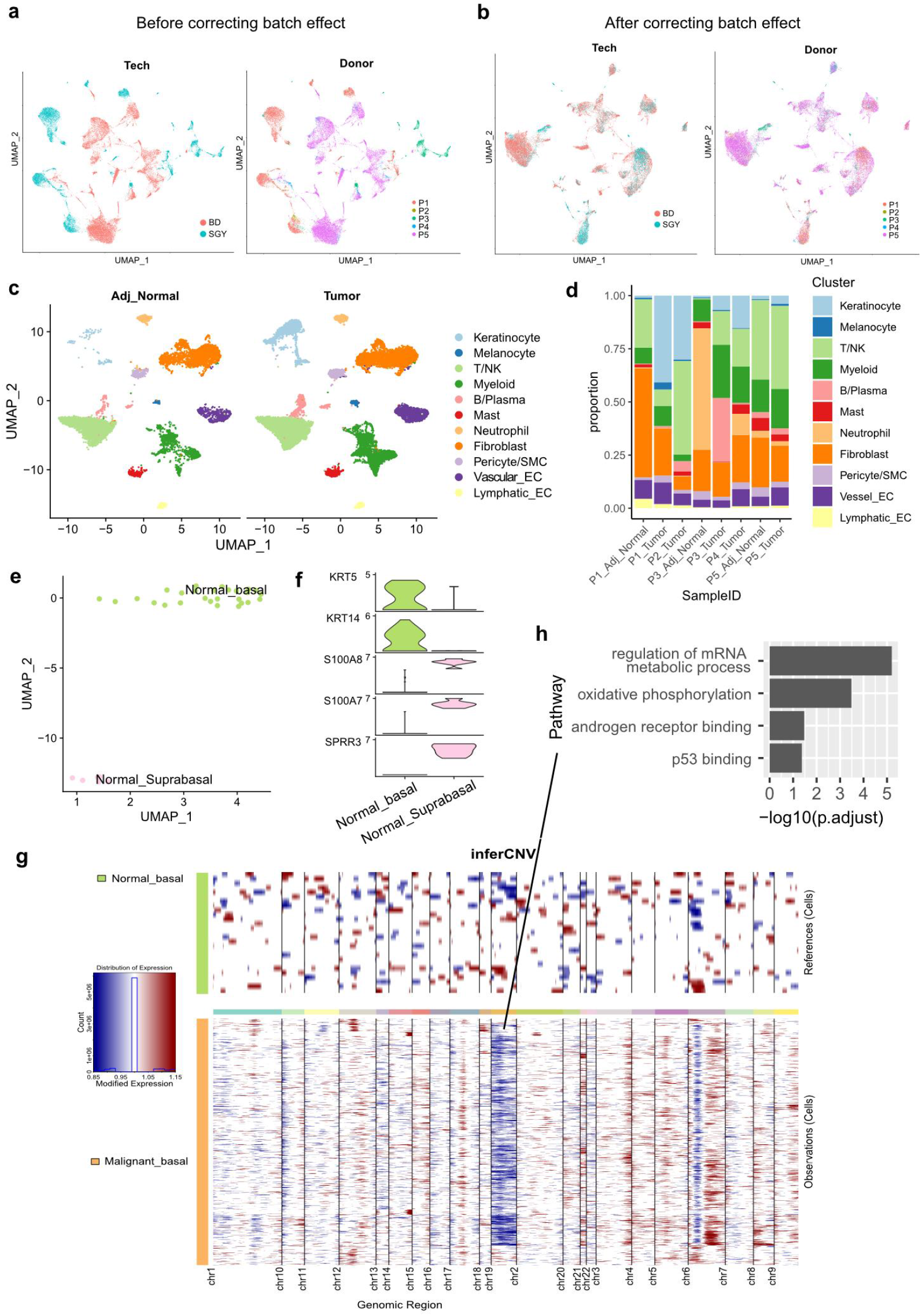
Single-cell atlas of human iBCC and identification of malignant basal cells. (a) The UMAP plot of cells before batch-correction on platforms and donors. (b) The UMAP plot of cells after batch-correction on platforms and donors. (c) The UMAP plot of major cell types from iBCC and ANS. (d) The bar chart showed the proportion of cell types in each sample. (e) The UMAP plot of keratinocytes sub-populations in ANS. (f) The expression of marker genes of keratinocytes sub-populations in ANS on Violin plot. (g) Heatmap showing large-scale CNVs of MBCs and NBCs. The normalized CNV levels were shown, Red indicated genomic amplifications and blue indicated genomic deletions. (h) The GO enrichment results based on genes of copy number loss on chromosome 19 in MBCs. CNV, copy number variation.

**Fig. S2.**
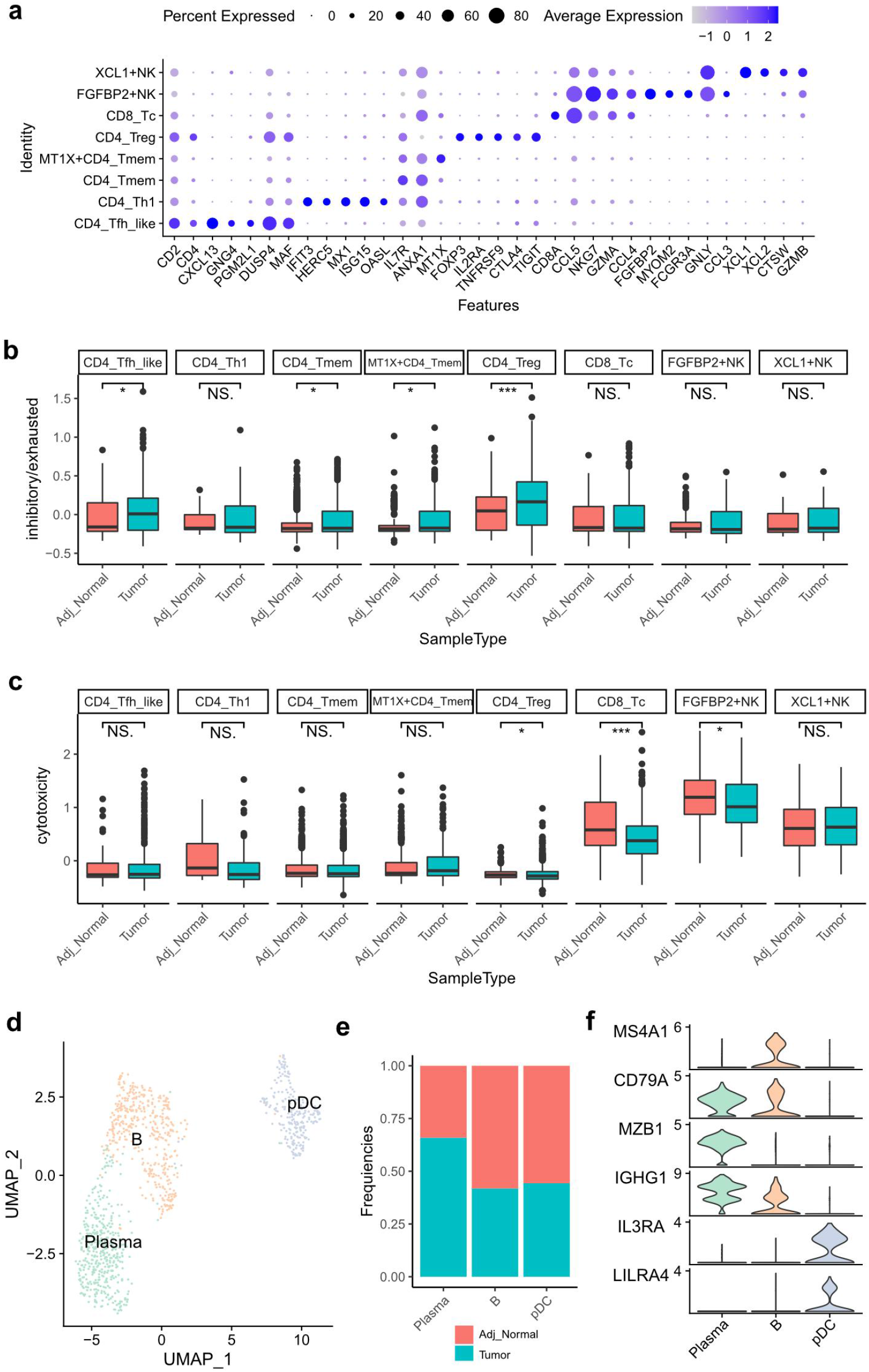
Cell subgroups of T/NK cells and B/Plasma cells. (a) Dot plot showing the marker genes of T/NK subgroups. (b) The inhibitory/exhausted scores among different cell types. (c) The cytotoxicity scores among different cell types. (d) The UMAP plot of B/Plasma compartment. (e) Frequencies of cells from ANS and iBCC present in each cell subgroup. f Violin plot showing the marker genes of B/Plasma subgroups. Wilcoxon signed-rank test, \**p* < 0.05, \*\**p* < 0.01, \*\*\**p* < 0.001. pDC, plasmatoid DC

**Fig. S3.**
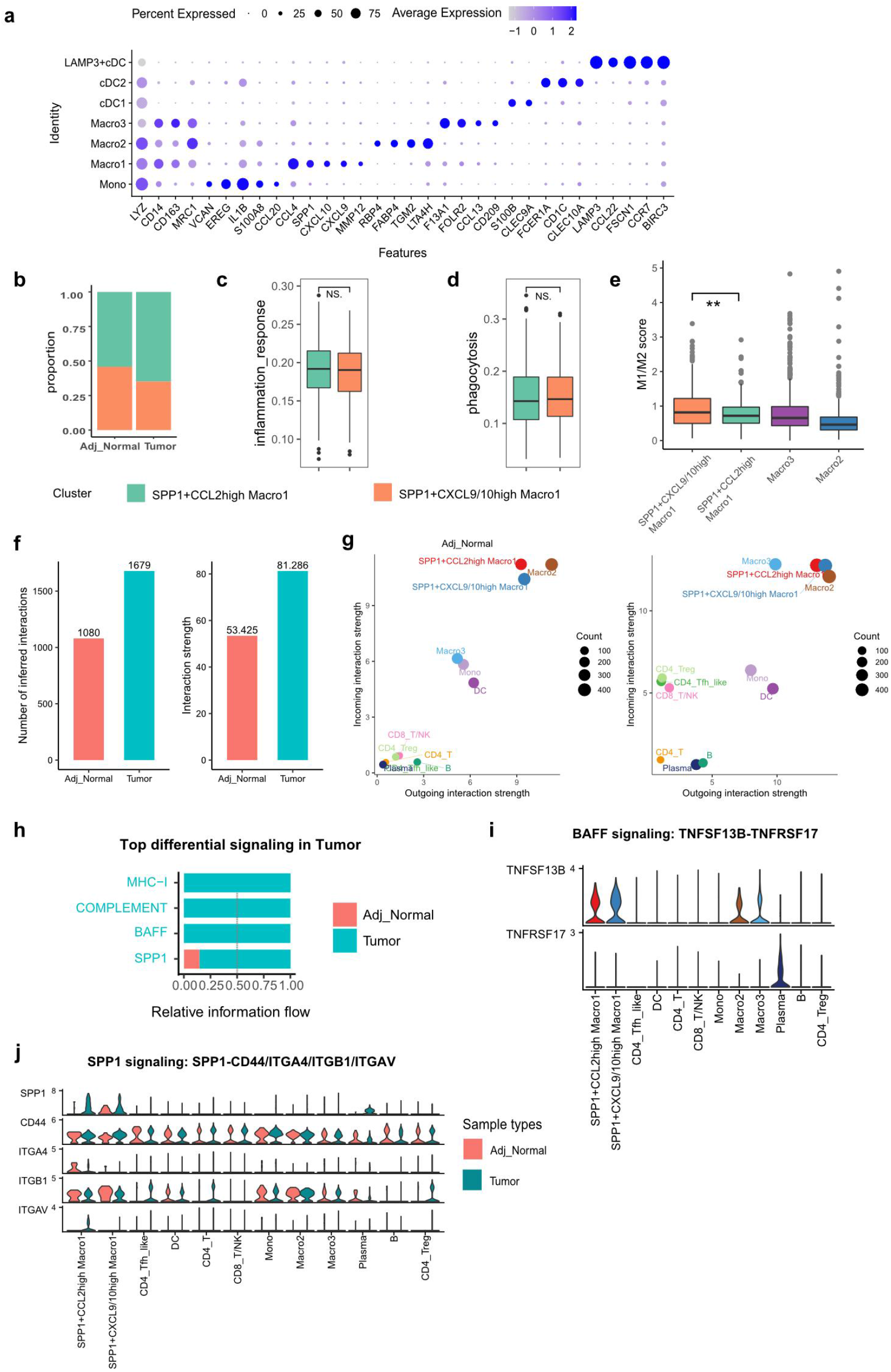
Heterogeneity of myeloid cells and cellular interactions among immune cells. (a) Dot plot showing the marker genes of myeloid subgroups. (b) The proportion of two subtypes of SPP1^+^ Macro1 in ANS and iBCC. (c, d) The GO terms scores similar in two subtypes of SPP1^+^ Macro1. (e) The M1/M2 score ratio among macrophage subtypes. (f) The number and weight of inferred interactions in ANS and iBCC. (g) The incoming and outgoing signaling strength of different cell types in ANS and iBCC. (h) The significant signaling pathways were ranked based on their differences in information flow between ANS and iBCC. We show the more enriched signaling pathways colored in blue in iBCC. (i) The expression of ligands and receptors among different cells in iBCC, and colors represent different cell types. (j) The expression of ligands and receptors among different cells, and colors represent different sample types.

**Fig. S4.**
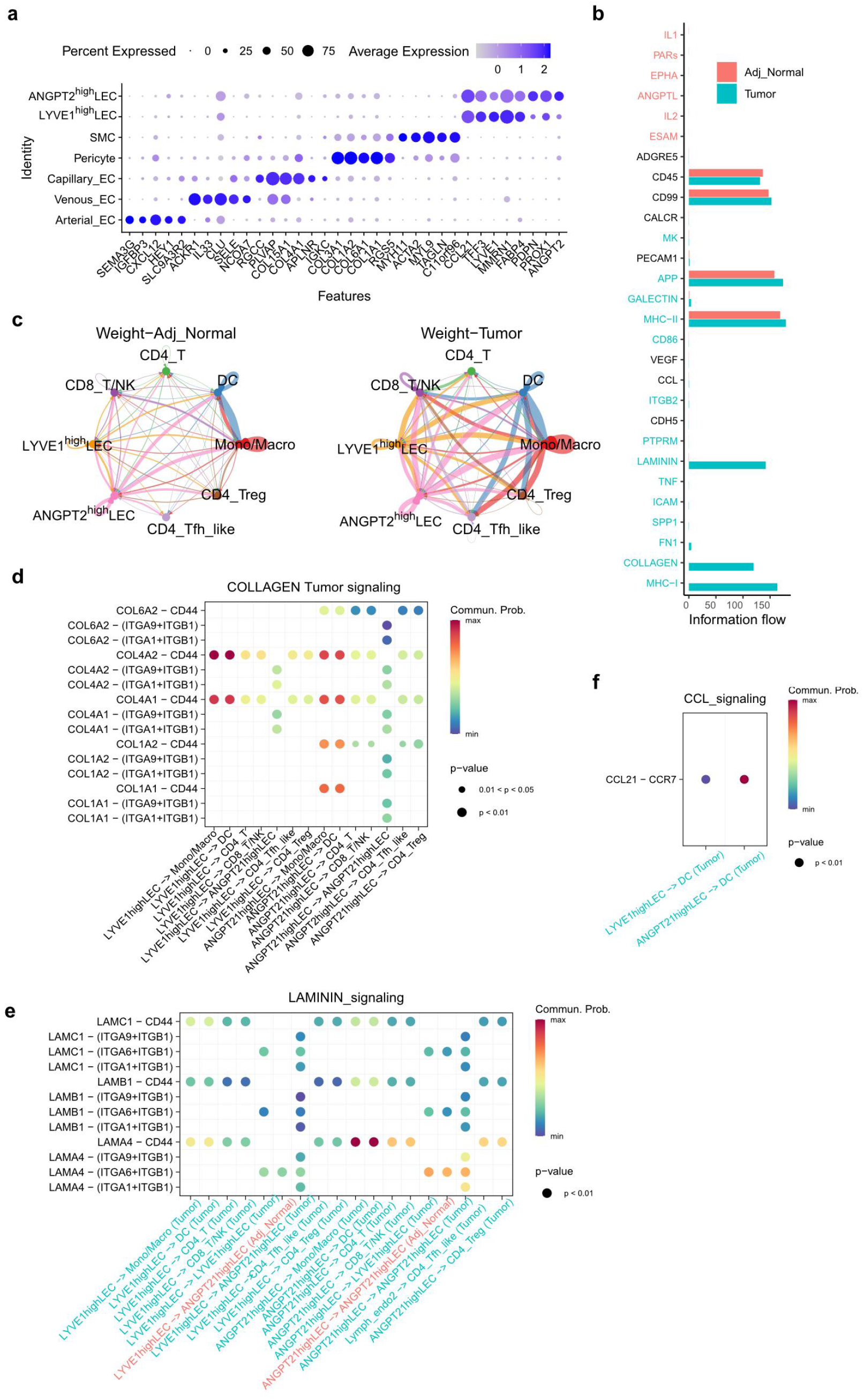
Tumor-associated ANGPT2^high^ lymphatic endothelial cells. (a) Dot plot showing the diffenrential expressed genes of mesenchymal subpopulations. (b) The significant signaling pathways were ranked based on their differences in overall information flow within the inferred networks between ANS and iBCC. (c) The weight of cell interactions across major immune cell types and LECs in ANS and iBCC. (d) Ligand–receptor interactions of COLLAGEN signaling pathway in iBCC. (e, f) Ligand–receptor interactions of LAMININ, CCL signaling pathway in ANS and iBCC. *p* value is represented by the size of each circle. The color gradient indicates the communication probability of interaction.

**Fig. S5.**
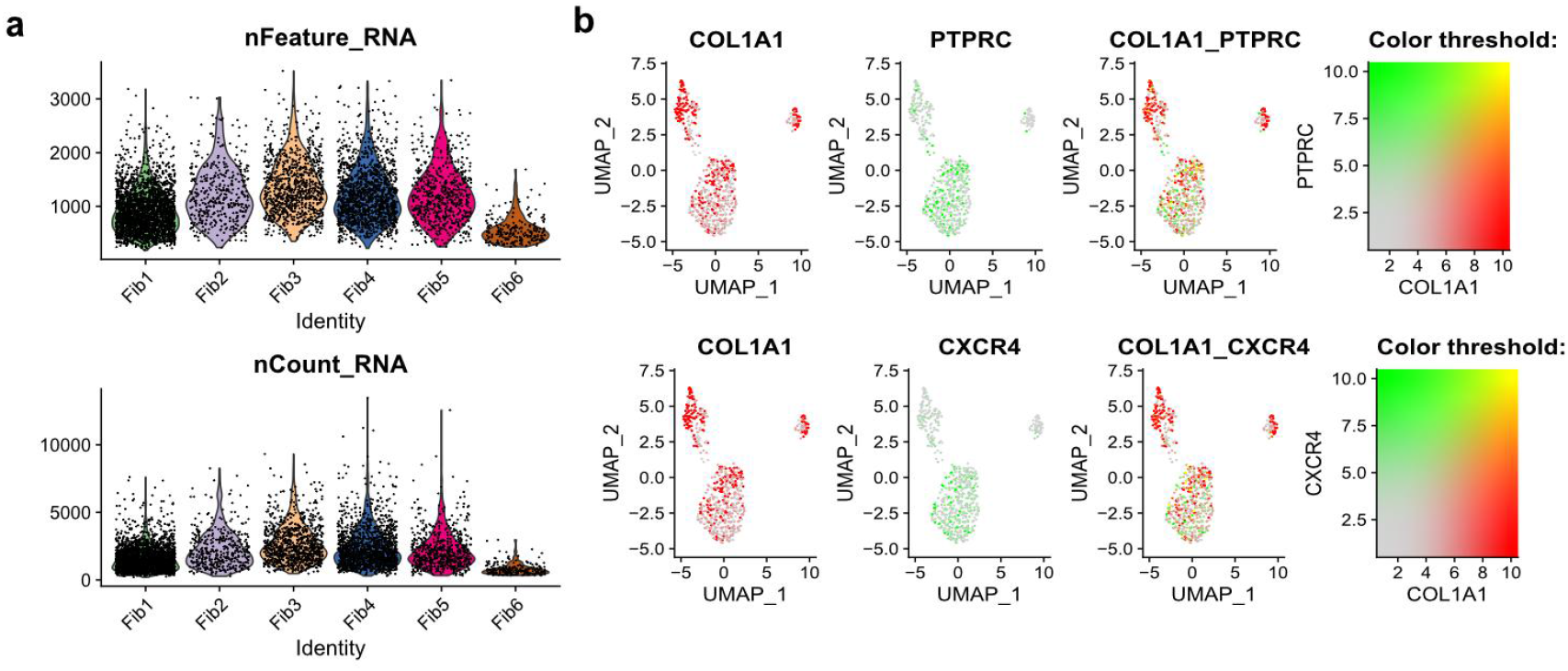
Identification of antigen-presenting fibroblasts. (a) The number of genes and counts of different fibroblast subgroups. (b) The split and merged expression level of two genes in Fib5 among UMAP. The colorization of yellow on cell dots signifying the co-expression level of the two genes.

**Fig. S6.**
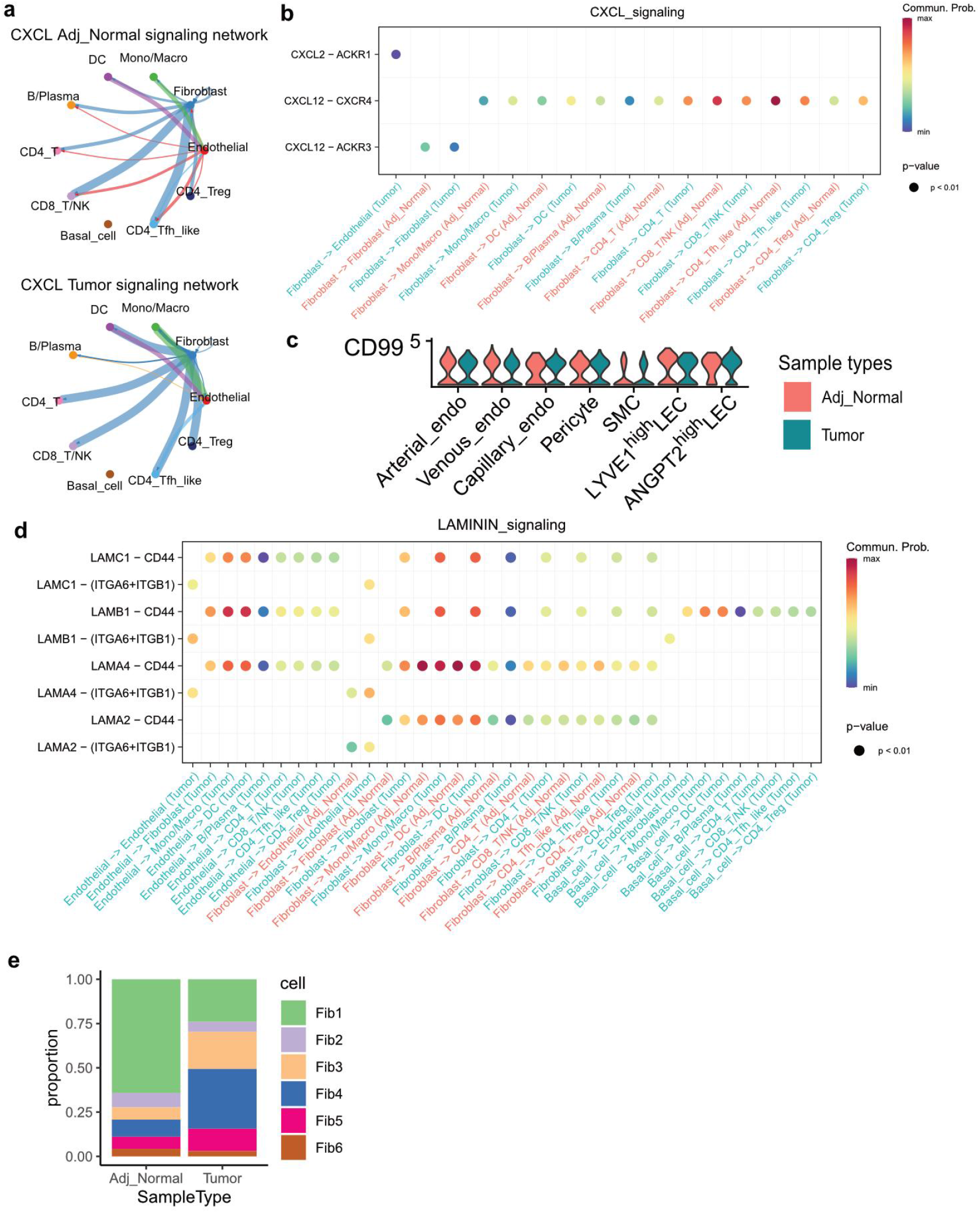
Globally cellular interactions in iBCC and ANS. (a) Cellular interactions of CXCL signaling pathway. (b) Ligand-receptor interaction probabilities of CXCL signaling sended from fibroblasts. (c) CD99 expression among endothelial cell subgroups. (d) Ligand-receptor interaction probabilities of LAMININ signaling. (e) The bar chart showed the proportion of fibroblast subgroups in ANS and iBCC.

**Fig. S7.**
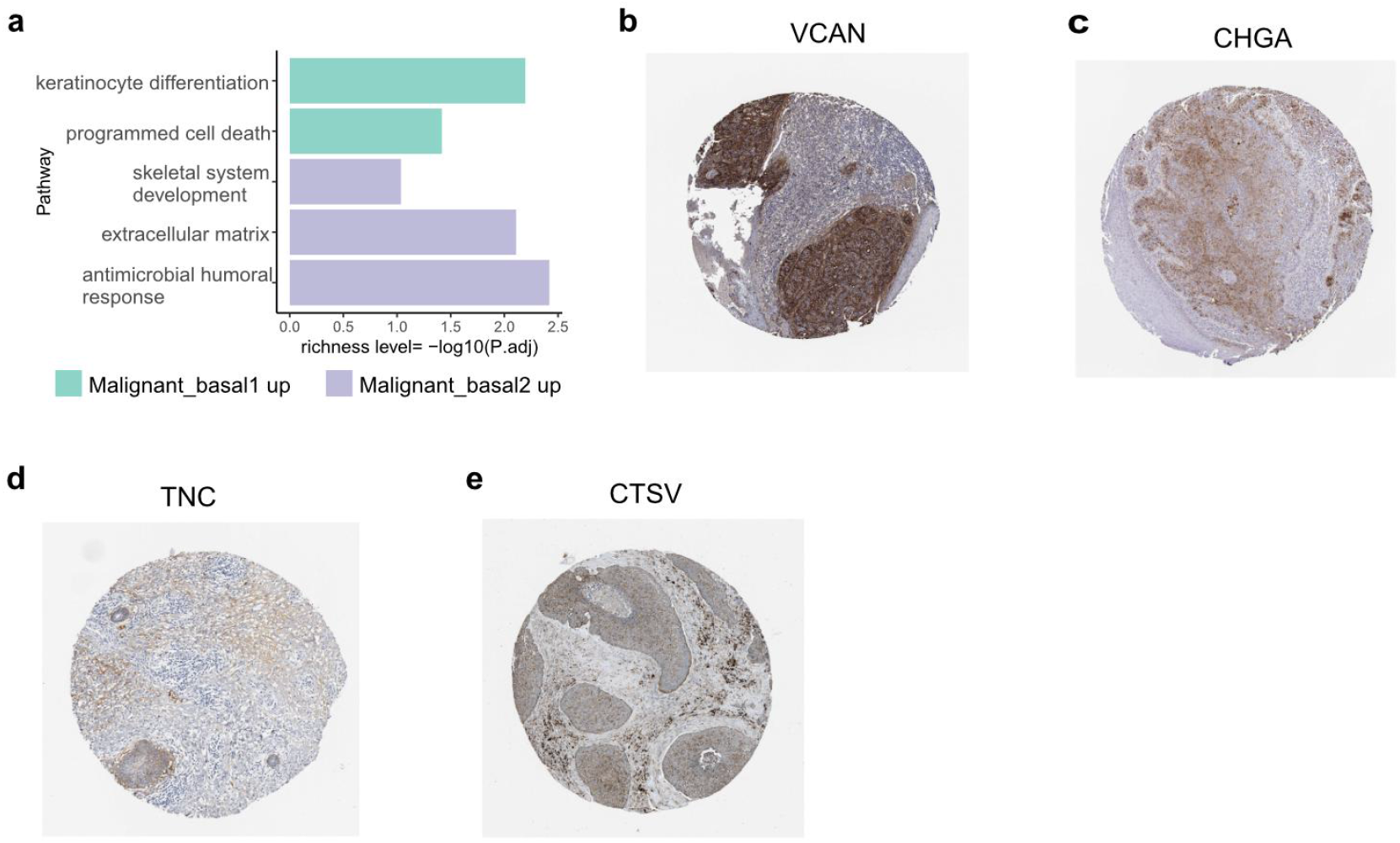
Two subtypes of malignant basal cells. (a) The enriched GO terms in MB1 versus MB2. (b-e) The IHC staining of VCAN, CHGA, TNC and CTSV in iBCC.

