## Supplementary Table 1 for "Single-cell profiling reveals the sustained immune infiltration, surveillance, and tumor heterogeneity of infiltrative BCC"

Table S1. The characteristics of the iBCC donors.

| Sample ID | Location | Sex | Tumor size | Platform |
| --- | --- | --- | --- | --- |
| P1-iBCC | Left lower eyelid | Female | 10*8mm | Singleron |
| P1-ANS | Adjacent normal skin | Female | - | Singleron |
| P2-iBCC | Right lower eyelid | Female | 2.0*1.5mm | Singleron |
| P3-iBCC | Right eyelid | Male | 2.5*1.5mm | Singleron |
| P3-ANS | Adjacent normal skin | Male |  | Singleron |
| P4-iBCC | Left lateral canthus | Male | 1.5*1.5mm | BD |
| P5-iBCC | Left lower eyelid | Male | 1.5*1.5mm | BD |
| P5-ANS | Adjacent normal skin | Male |  | BD |
