## Supplementary Table 2 for "Single-cell profiling reveals the sustained immune infiltration, surveillance, and tumor heterogeneity of infiltrative BCC"

Table S2. Cell populations of major cell type in each sample.

|  | P1_Adj_  Normal | P1_  Tumor | P2_  Tumor | P3_Adj_  Normal | P3_  Tumor | P4_  Tumor | P5_Adj_  Normal | P5_  Tumor | sum |
| --- | --- | --- | --- | --- | --- | --- | --- | --- | --- |
| B/Plasma cell | 13 | 34 | 14 | 2 | 517 | 9 | 124 | 373 | 1086 |
| Fibroblast | 2040 | 1009 | 18 | 87 | 279 | 268 | 1055 | 2132 | 6888 |
| Keratinocyte | 40 | 1851 | 87 | 4 | 116 | 184 | 77 | 489 | 2848 |
| Lymphatic_EC | 177 | 90 | 4 | 2 | 4 | 11 | 45 | 154 | 487 |
| Mast cell | 61 | 6 | 6 | 13 | 5 | 54 | 274 | 403 | 822 |
| Melanocyte | 29 | 154 | 2 | 2 | 10 | 4 | 19 | 128 | 348 |
| Myeloid cell | 299 | 430 | 9 | 46 | 431 | 207 | 695 | 2343 | 4460 |
| Neutrophil | 16 | 11 | 1 | 255 | 1 | 119 | 146 | 265 | 814 |
| Pericyte/SMC | 49 | 144 | 5 | 18 | 28 | 39 | 199 | 343 | 825 |
| T/NK | 906 | 346 | 127 | 2 | 272 | 213 | 1686 | 4921 | 8473 |
| Vascular_EC | 352 | 459 | 16 | 16 | 60 | 97 | 201 | 1082 | 2283 |
| sum | 3982 | 4534 | 289 | 447 | 1723 | 1205 | 4521 | 12633 | 29334 |
