## Supplementary Table 3 for "Single-cell profiling reveals the sustained immune infiltration, surveillance, and tumor heterogeneity of infiltrative BCC"

Table S3. Immune associated gene sets.

| Name | Genes |
| --- | --- |
| cytotoxicity | PRF1, IFNG, GNLY, NKG7, GZMB, GZMA, GZMH, KLRK1, KLRB1, KLRD1, CTSW, CST7, CCL4, CCL5 |
| Inhibitory/exhausted | PDCD1, HAVCR2, LAG3, TIGIT, CTLA4, ENTPD1, LAYN, EBI3, CD96 |
| M1 | IL23A, IL12A, IL12B, NOS2, SOCS3, TNF, CXCL9, CXCL10, CD86, IL1A, IL1B, IL6, CCL5, IRF5, IRF1, CD40, IDO1, KYNU, CCR7 |
| M2 | IL4R, CCL4, CCL13, CCL20, CCL17, CCL18, CCL22, CCL24, LYVE1, VEGFA, VEGFB, VEGFC, VEGFD, EGF, CTSA, CTSB, CTSD, TGFB1, TGFB2, TGFB3, MMP14, MMP19, MMP9, CLEC7A, WNT7B, TNFSF12, TNFSF8, CD276, VTCN1, MSR1, FN1, IRF4 |
