## Supplementary Table 4 for "Single-cell profiling reveals the sustained immune infiltration, surveillance, and tumor heterogeneity of infiltrative BCC"

Table S4. Demographic Characteristics of Patients With iBCC

|  | MDK expression | |  |  |
| --- | --- | --- | --- | --- |
| Characteristics | Low (≤2) | High (>2) | No. of patient (N=39) | *p* |
| **Age (years)** |  |  |  | 0.6927 |
| <60 | 6 | 4 | 10 |  |
| ≥60 | 21 | 8 | 29 |  |
| **Gender** |  |  |  | 0.9999 |
| Male | 16 | 6 | 22 |  |
| Female | 12 | 5 | 17 |  |
| **Duration of the tumor** |  |  |  | 0.4801 |
| <5 years | 13 | 7 | 20 |  |
| ≥5 years | 15 | 4 | 19 |  |
| **Ulcer** |  |  |  | 0.9999 |
| Yes | 23 | 9 | 32 |  |
| No | 5 | 2 | 7 |  |
| **Recurrence** |  |  |  | 0.6927 |
| Yes | 6 | 3 | 9 |  |
| No | 22 | 8 | 30 |  |
| **Depth of infiltration (mm)** |  |  |  | 0.0047 |
| <5 | 23 | 3 | 26 |  |
| ≥5 | 5 | 7 | 13 |  |
