## Supplementary Table 5 for "Single-cell profiling reveals the sustained immune infiltration, surveillance, and tumor heterogeneity of infiltrative BCC"

Table S5. Detailed information of iBCC patients.

| **Number** | **MDK expression** | **Infiltration Depth** | **Duration (years)** | **Ulcer** | **Recurrence** |
| --- | --- | --- | --- | --- | --- |
| 1 | 3 | 7.00 |  | YES | YES |
| 2 | 1 | 1.00 | 10 | NO | NO |
| 3 | 2 | 2.00 | 2 | YES | NO |
| 4 | 1 | 2.00 | 4 | YES | NO |
| 5 | 3 | 4.00 | 0.5 | YES | NO |
| 6 | 2 | 5.00 | 6 | YES | NO |
| 7 | 2 | 1.20 | 8 | NO | NO |
| 8 | 3 | 5.00 | 4 | YES | NO |
| 9 | 3 | 8.00 | 8 | YES | NO |
| 10 | 2 | 2.50 | 10 | YES | NO |
| 11 | 1 | 1.60 | 5 | YES | YES |
| 12 | 3 | 10.00 | 8 | YES | YES |
| 13 | 1 | 5.50 | 1 | YES | YES |
| 14 | 3 | 1.50 | 4 | YES | NO |
| 15 | 2 | 4.00 | 10 | YES | NO |
| 16 | 1 | 2.00 | 0.5 | YES | NO |
| 17 | 2 | 1.20 | 5 | NO | NO |
| 18 | 1 | 1.90 | 4 | YES | NO |
| 19 | 2 | 4.00 | 1 | YES | NO |
| 20 | 1 | 3.00 | 7 | YES | YES |
| 21 | 1 | 1.30 | 0.42 | YES | NO |
| 22 | 2 | 2.50 | 10 | YES | NO |
| 23 | 4 | 2.50 | 7 | YES | YES |
| 24 | 1 | 2.50 | 0.5 | YES | NO |
| 25 | 1 | 1.00 | 6 | YES | NO |
| 26 | 2 | 7.00 | 7 | YES | YES |
| 27 | 1 | 10.80 | 10 | YES | YES |
| 28 | 4 | 5.00 | 4 | NO | NO |
| 29 | 3 | 8.00 | 1 | YES | NO |
| 30 | 2 | 7.00 | 10 | YES | NO |
| 31 | 4 | 5.00 | 2 | NO | NO |
| 32 | 2 | 3.00 | 5 | YES | NO |
| 33 | 1 | 2.50 | 2 | YES | NO |
| 34 | 0 | 1.50 | 2 | NO | NO |
| 35 | 1 | 1.00 | 1 | YES | NO |
| 36 | 1 | 3.00 | 9 | YES | NO |
| 37 | 1 | 2.00 | 3 | YES | NO |
| 38 | 3 | 7.00 | 2 | YES | NO |
| 39 | 2 |  | 4 | NO | YES |
